# Uncovering Social States in Healthy and Clinical Populations Using Digital Phenotyping and Hidden Markov Models: Observational Study

**DOI:** 10.1101/2024.06.07.24308598

**Authors:** Imogen E. Leaning, Andrea Costanzo, Raj Jagesar, Lianne M. Reus, Pieter Jelle Visser, Martien J.H. Kas, Christian Beckmann, Henricus G. Ruhé, Andre F. Marquand

## Abstract

**Background:** Brain related disorders are characterized by observable behavioral symptoms, for example social withdrawal. Smartphones can passively collect behavioral data reflecting digital activities, such as communication app usage and calls. This data is collected objectively in real time, avoiding recall bias, and may therefore be a useful tool for measuring behaviors related to social functioning. Despite promising clinical utility, analyzing smartphone data is challenging as datasets often include a range of missingness-prone temporal features.

**Objective:** Hidden Markov Models (HMMs) provide interpretable, lower-dimensional temporal representations of data, allowing missingness. We aimed to investigate the HMM as a method for modeling smartphone time series data.

**Methods:** We applied an HMM to an aggregate dataset of smartphone measures designed to assess phone-related social functioning in healthy controls (HCs), participants with schizophrenia, Alzheimer’s disease (AD) and memory complaints. We trained the HMM on a subset of HCs (n=91) and selected a model with socially “active” and “inactive” states, then generated hidden state sequences per participant and calculated their “total dwell time”, i.e. the percentage of time spent in the socially active state. Linear regression models were used to compare the total dwell time to social and clinical measures in a subset of participants with available measures, and logistic regression was used to compare total dwell times between diagnostic groups and HCs. We primarily report results from a two-state HMM but also verified results in HMMs with more hidden states, and trained on the whole participant dataset.

**Results:** We identified lower total dwell times in AD (n=26) versus withheld HCs (n=156) (odds ratio=0.95, FDR corrected *P*<.001), as well as in participants with memory complaints (n=57) (odds ratio=0.97, FDR corrected *P*=0.004). The AD result was very robust across HMM variations, whilst the memory complaints result was less robust. We also observed an interaction between AD group and total dwell time when predicting social functioning (FDR corrected *P*=0.02). No significant relationships regarding total dwell time were identified for participants with schizophrenia (n=18).

**Conclusions:** We found the HMM to be a practical, interpretable method for digital phenotyping analysis, providing an objective phenotype that is a possible indicator of social functioning.

## Introduction

Many psychiatric and neurological diseases exhibit observable behaviors that are indicative of the underlying condition. For example, social functioning is negatively impacted in a broad range of conditions, including schizophrenia (SZ), Major Depressive Disorder (MDD), anxiety disorders and Alzheimer’s disease (AD) (e.g. [1–3]), often cumulating in social withdrawal. Social withdrawal, indicated by reduced social interaction[1], can be observed as people engage less with those around them. However, successfully measuring behavioral components such as social withdrawal is challenging, as reports of behavior are subjective and susceptible to recall bias, with questionnaires often being burdensome to complete. There is therefore a need to develop practical, objective tools to monitor these symptoms, for example to predict or measure clinically relevant changes.

The field of digital phenotyping is developing to meet such a need. Digital phenotyping involves the development of behavioral or physiological markers calculated from digital measures. “Digital phenotype” is a broad term referring to a quantified digital behavior (such as use of smartphone applications (apps)) or behavior measured using a digital signal (such as movement measured using GPS). These measures avoid issues of recall bias as they are objective and can be acquired in real-time as participants go about their day, also meaning they have high ecological validity. A popular tool to collect digital phenotyping data is the smartphone. Given the highly common place of smartphones in society, the smartphone is convenient as it does not require participants to change their behavior or routines; a monitoring application, for example “Behapp”,[4] “Mood mirror”[5] or “RADAR-base pRMT”,[6] can be installed on their own phone and run passively in the background to collect data, without user intervention.

Modern smartphones have a large number of sensors and functionalities, including various apps, calling capabilities, WiFi, GPS, accelerometer and Bluetooth, which can be leveraged to model different aspects of behavior (including social contacts, movement patterns and app usage (e.g. [7,8])). Many of these datastreams are direct measures of digital behaviors that can be used as proxy measures of social behavior, for example the use of communication apps could indicate how connected someone is with their contacts. Whilst using these measures requires inferences to be made about behavior, their objective nature and the range in measures means they are a promising tool for modeling social behavior.

Moreover, there are many ways in which these data can be processed. For example, the duration, circadian rhythm or statistical measures can be calculated (such as mean and standard deviation of a behavior across time) or the occurrences of the behavior counted.[9] This often leads to datasets with many features reflecting various different smartphone-measured behaviors. A major problem affecting digital phenotyping is that platforms are often prone to missing data due to the difficulties of real-world longitudinal data collection, leading to missing values across all or a subset of these features.[9]

The issues and complexities observed in digital phenotyping research give rise to multiple analytic challenges: processing the collected feature sets, often representing a wide range of seemingly distinct observed behaviors with potentially similar underlying causes, requires many model decisions. Appropriate methods are therefore needed to analyse this multi-faceted data containing missingness, in order to produce meaningful, lower-dimensional data representations. These representations may be more usable and informative about the underlying behavioral states of participants relative to the individual features. Models should also aim to be interpretable by not only researchers but also clinicians (and patients), to facilitate their use in clinical practice. A further property that would enable their use in this context is that they can preserve the time domain, as one of the goals of smartphone digital phenotyping is to be able to make useful clinical predictions that can enable early intervention. Many digital phenotyping studies have focused on time-averaged features and analyses, and a shift towards more direct investigations of temporal dynamics is expected to improve clinical utility.[9]

Additionally, given the range in symptomatologies experienced by people within many neuropsychiatric disorders, it may be useful to define a reference distribution that could represent a “standard operating range” for a given population or participant, where deviations from this range can then be conceptualized as potentially signaling transitions into different behavioral modes of functioning, as is done in normative modeling[10,11] or anomaly detection applications.[12,13] This reference distribution could be, for example, data from healthy controls (HCs) or from periods when individuals are not currently experiencing a relapse of their disorder. This approach may also help to leverage more easily collectable periods of data, as it can be challenging to capture periods containing relapses or the symptom severity range that is of interest, leading to smaller volumes of data for these periods.

Currently, digital phenotyping studies employ a broad range of modeling approaches. For example, investigating associations between neuropsychiatric symptoms and summary measures (e.g. total number of places visited, mean duration of communication app usage),[14] clustering of digital phenotypes to investigate transdiagnostic symptom classification,[15] linear mixed effects models accounting for repeated measures of time-averaged features ([16–19]), multivariate anomaly detection to identify relapse in SZ[20] and joinpoint regression to identify changes in the trajectory of digital phenotypes (e.g. step count).[21]

In this study we propose the use of a Hidden Markov Model (HMM) (e.g. [22]) as a method to model digital phenotyping time series data. This provides several appealing features, namely that HMMs (1) can meaningfully combine different behavioral features, (2) reflect changes in behavior over time, (3) provide readily interpretable summary statistics and (4) naturally accommodate missingness. HMMs provide interpretable, lower dimensional representations of the data using latent (i.e. hidden) states, where the observed time series channels are represented as a sequence of these hidden states. Each hidden state has associated “emission probabilities” indicating the probability that a set of observed behaviors occur when the sequence is in said hidden state, allowing for informative behavioral states to be derived by representing more than one feature per state. Changes in behavior through time are modeled via transitions between these hidden states. Importantly for digital phenotyping, HMMs contain intrinsic mechanisms for handling missing data. HMMs have been used in many applications for modeling behavior, for example to model drinking patterns in people with an alcohol use disorder,[23] cocaine dependence,[24] sleep patterns represented in neuroimaging data,[25] mobility data (e.g. [26,27]), weekly psychotic depressive symptom profiles,[28] weekly depressive symptom profiles,[29] and actigraphy and survey data reflecting behavior and affect in college students.[30]

While our approach is widely applicable to digital phenotyping time series, in this work we demonstrate its application to data collected using the Behapp monitoring application,[31] which collects passive data related to app usage, calls, GPS, WiFi and overall phone usage, reflecting the periods the phone was unlocked. We applied an HMM to a combined dataset of phone usage and communication-related features from participants in the “Psychiatric Ratings using Intermediate Stratified Markers” (PRISM)[32] and Hersenonderzoek (HO)[14] studies, demonstrating how an HMM can successfully represent digital phenotyping time series. The model was initially trained on a set of HCs with low missingness to provide a high-quality dataset for training, which was treated as a “reference category”. The trained model was then applied to HCs with higher missingness, and participants with AD, SZ and healthy participants with memory complaints (“subjective cognitive complaints”; SCC), to investigate the applicability of such a model to clinical groups and participants with lower data availability.

Hidden state sequences were generated for these participants, and we then calculated a digital phenotype derived from the HMM for each participant, namely the “total dwell time”. Rather than being a directly observed digital phenotype (such as the percentage of time spent using communication apps), the total dwell time provides the percentage of time the participant spent in a hidden behavioral state derived from the observed digital measures. This digital phenotype was then linked to clinical measures including diagnostic group and social functioning, demonstrating the clinical value of this approach.

## Method

### Participants

This analysis utilized data from participants from the PRISM and HO studies. We chose to combine these datasets in our analysis due to the overlap in populations, as both studies included participants with AD and consequently, similarly age-matched HCs, meaning we could have an increased sample size for the AD and HC groups.

#### PRISM

The PRISM study aims to investigate social withdrawal in two brain disorders, SZ and probable AD ([32,33]). Participants with AD, SZ, and age– and gender-matched HCs were recruited across centres in Spain (Hospital General Universitario Gregorio Marañón and Hospital Universitario de La Princesa, in Madrid) and the Netherlands (University Medical Center Utrecht, Leiden University Medical Center and Amsterdam UMC, location VUmc).

Participants with SZ were required to be within the age range of 18-45 years (inclusive), and to have a DSM-IV (Diagnostic and Statistical Manual of Mental Disorders) diagnosis of SZ confirmed by the Mini-International Neuropsychiatric Interview (MINI). Participants were required to have experienced at least one psychotic episode, to have had a maximum disease duration of 10 years since diagnosis, and for any antipsychotic medication dosage to have been stable for a minimum of 8 weeks. As PRISM aimed to investigate social withdrawal linked with negative symptoms (and not as a consequence of other sources such as psychosis), participants with SZ were excluded if they rated highly for positive symptoms (≥22 on the positive symptom factor of the 7-item Positive and Negative Syndrome Scale (PANSS)[34]). A positive symptom indicates an additional experience an individual is having, such as a hallucination or delusion, as opposed to a negative symptom which indicates a deficit in an already existing function, such as a deficit in concentration. Whilst SZ is commonly associated with positive symptoms, negative symptoms also form a large component of the disorder. Participants with AD were required to be within the age range of 50-80 years, to meet the classification of “Probable AD” based on the National Institute on Aging and the Alzheimer’s Association (NIAAA) criteria, and to have a Mini-Mental State Examination (MMSE)[35] score of 20-26. For both participants with SZ and AD, it was required that participants were not socially withdrawn due to other reasons such as their external circumstances, a comorbid medical disorder or disability. These factors were evaluated during the intake interview.

HCs were recruited in the age ranges of 18-45 and 50-80, and were required to have an approximately average MMSE score according to their age and years of education. Participants were excluded if they met the criteria for an Axis-I psychiatric disorder (assessed by the MINI), or a neurological disease associated with cognitive impairment. For further details of inclusion/exclusion criteria for all participant groups see the PRISM study overview.[32]

In addition to Behapp data collection, measures of clinical and social functioning were acquired. The self-report Social Functioning Scale (SFS)[36] and the De Jong Gierveld Loneliness and Affiliation Scale[37] were administered to all participants, the MMSE was administered to HCs and participants with AD, and the PANSS was administered to participants with SZ.

#### Hersenonderzoek

Participants with probable AD, “Subjective cognitive complaints” (SCC) and age-matched HCs were recruited across the Netherlands by The Dutch Brain Research Registry (Hersenonderzoek.nl), providing demographics and health-related information online via the Hersenonderzoek.nl platform.[14] Participants indicated the presence of probable AD. To classify participants as SCC or HC respectively, participants indicated the absence of neurological or psychiatric diseases with or without memory complaints. The minimum age for inclusion was 45 years.

### Ethical Considerations

For the participating research centers in the Netherlands, PRISM was approved by the Ethical Review Board University Medical Centre of Utrecht (17-021/D). For the participating research centers in Spain, PRISM was approved by the Comité Ético de Investigación Clínica Hospital General Universitario Gregorio Marañón (59359). PRISM participants were deemed by the researcher and caregivers to be sufficiently competent to participate in the study. Approval for HO was provided by the Ethical Review Board VU University Medical Centre (2017.254). All PRISM and HO participants provided informed consent before participation commenced. In the PRISM study, participants received both travel expenses and compensation for their time. For the HO study, it was possible to receive travel expenses. In both studies participants’ data was de-identified. Participants could request the deletion of their collected data from the database at any time, in line with the GDPR.

### Behapp Acquisition

The smartphone application “Behapp”[31] was installed on participants’ smartphones. Behapp passively collected smartphone-usage data for a period of 42 days without storing any content of messages and calls, in compliance with the European Privacy Regulation.[38] The classification of each app used by participants was gathered from the Google Play Store, so that apps could be grouped by type, including social media and communication apps. During the time of data collection (PRISM: August 2017 – May 2019; HO: March 2018 – January 2020), Behapp was only available on Android smartphones, and so PRISM participants who did not have their own Android smartphone were supplied with one for the duration of study participation. However, this was not done for HO participants in accordance with the study design, and only two PRISM participants used a study-provided phone. For each activity (e.g. use of an app), the respective start and end timestamps were stored.

### Preprocessing

#### Smartphone Channels

Phone usage was split into five categories, referred to as “channels”: social media app usage, communication app usage, incoming calls, outgoing calls, and overall phone usage. GPS channels were also available. Since many of these measures are sparsely sampled, each channel was aggregated into hourly bins, and the percentage of each hour for which each activity was carried out was calculated. For example, a participant may spend 100% of an hour using their phone, 50% on social media, 40% on communication apps, 0% making/receiving calls and 10% using another functionality such as Google Maps. Even with the temporal resampling, many of these phenotypes have highly zero-inflated distributions (see Figure S1 in Multimedia Appendix 1), which can be difficult to handle natively. Therefore, for each hourly timepoint, these percentages were grouped into discrete bins instead of continuous percentages: binary bins reflecting either no or some activity carried out in the hour (0% activity; >0 activity). We chose this low threshold to define activity as many of the activities we investigate may still be meaningful despite their short duration, for example the time it takes to send a message. We have conducted a sensitivity analysis to understand the impact that this activity threshold has, finding that a threshold requiring an activity to be carried out for at least 5% of an hour provides comparable results to the HMMs presented here, however this was no longer the case for a 10% threshold. With this threshold very few hours are classified as containing activity (see Figure S1 in Multimedia Appendix 1).

Digital phenotyping data is prone to missingness. Therefore, we developed two measures to identify whether data had been successfully collected by Behapp for each hour, with one measure reflecting overall data availability and the other reflecting data availability specific to GPS (a sensor which is especially prone to missingness). These measures were required so that we could differentiate between values that were zero because a participant was not using their phone and values that were zero because data was not successfully collected. These measures capitalize on the sampling frequency of the location and other data sources like WiFi data (which are both independent of active phone usage); as this frequency is expected to be greater than once per hour, a frequency lower than one sample per hour in the location data indicates missing location data, and a frequency lower than one sample per hour in all types of data (including WiFi) indicates that overall data was not being collected successfully. Therefore, one of these measures reflected overall data availability, and the other measure was specific to GPS data availability. The distributions for these measures are provided in Figures S2-S5 in Multimedia Appendix 1. Due to low GPS data availability acquired using the version of Behapp used in these studies, it was decided not to include the GPS channels in this analysis. Therefore, any missingness that occurred in the included channels occurred across all channels at the same timepoints (i.e. it is not possible to have data missing at a timepoint in, for example, only the social media channel and not the other channels).

To account for any changes in behavior that may have arisen from study onboarding (i.e. participant attending assessments at study location), the first day of each participant’s Behapp data was excluded. As a consequence, all time series began at midnight. If the overall data availability measure indicated missing data, then the channels were marked as “NA”. Since missing data are handled natively by the HMM implementation we employed,[22] as explained below, no missing data imputation was carried out on the data.

#### Division into Training and Validation Sets Based on Missing Data and Diagnostic Group

Participants were split into training and validation sets, with the training set used to train the model and the validation set used to investigate relationships between HMM-derived digital phenotypes and clinical measures. All participants with SZ, AD or SCC were assigned to the validation set (as well as a subset of withheld HCs), so that the HMM could be trained on HCs, akin to training on a reference category.[10] To ensure that the HMM was trained on high quality data (i.e. time series with low levels of missingness), HCs meeting an overall data availability criterion of at least 90% of timepoints available across their time series were assigned to the training set. No minimum requirement was set for Behapp participation length, so that shorter time series that did not have missingness issues during data collection were still included. We randomly selected 15 of these high data availability HCs and retained them in the validation set, to allow for some amount of data availability matching between HCs in the training and validation sets, also increasing the number of HCs in the validation set with social and clinical scale measures available. The distributions of time series lengths for training and validation participants are provided in Figures S6 and S7, and distributions of data availability are provided in Figures S2-S5 in Multimedia Appendix 1. Additionally, we investigated equivalent HMMs trained on the entire dataset (i.e. no training/validation split) for insight into how the dataset split was affecting the learned model.

### Overview of Hidden Markov Model

An overview of the main approach used in this study can be seen in Figure 1. The HMM models the observed smartphone data channels using a smaller number of hidden states, where each hidden state has corresponding probable values in these observed channels. Through time the participant then switches between different hidden states. Mathematically, given a sequence of observed variables **x**_*t*_ and hidden states **z**_*t*_, at time *t* = 1, …, *T*, the joint distribution for this model can be specified as:

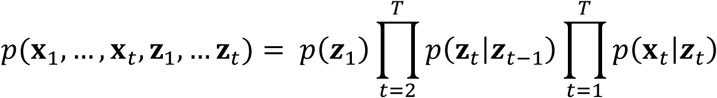

**Figure 1:**
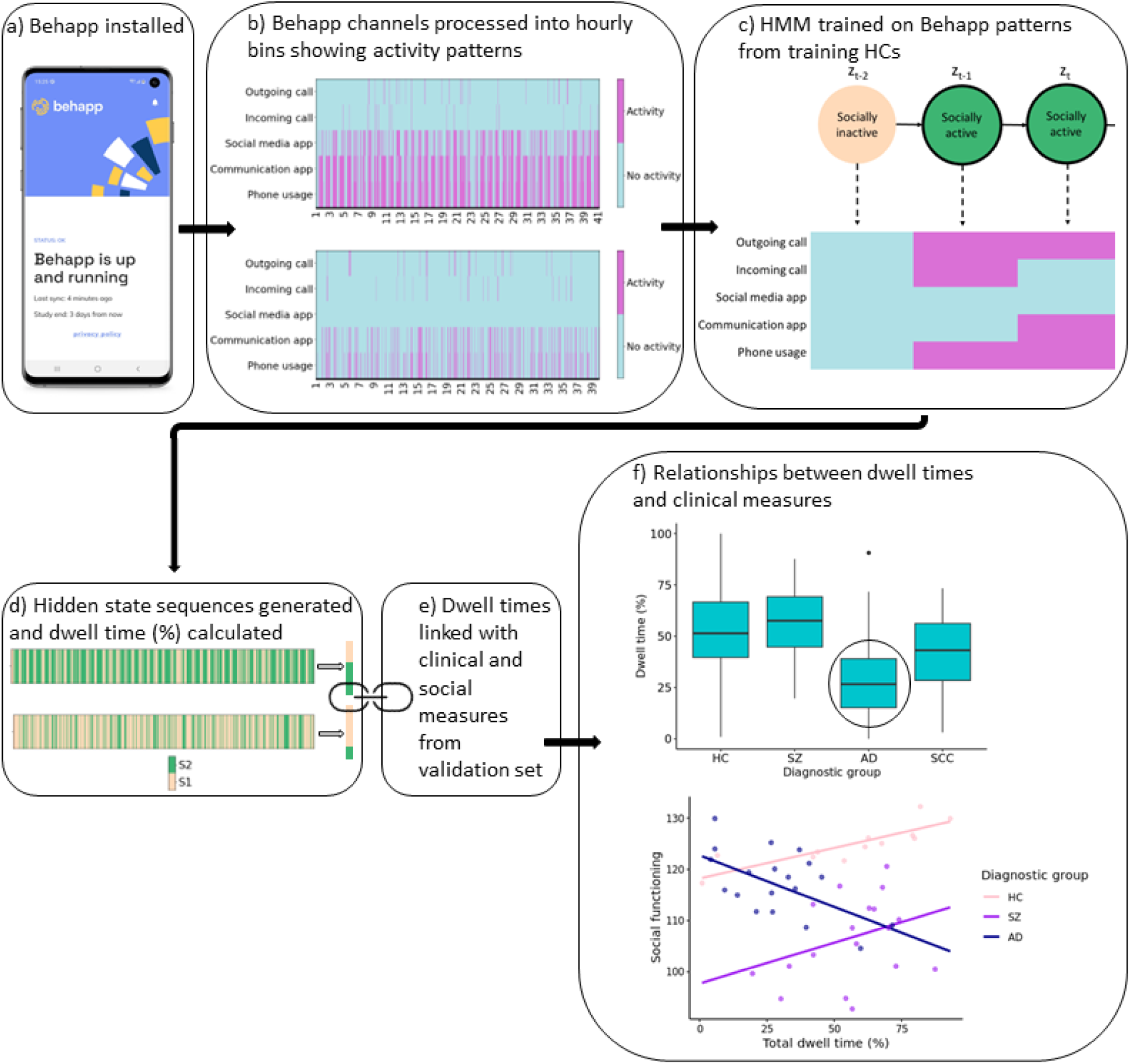
Overview of Hidden Markov Model approach: this flowchart highlights the main processing and modeling steps involved in the HMM method. a) Upon participation commencement the Behapp application was installed and collected data passively. b) This data was processed into bins that showed whether an activity was being carried out each hour. c) The HMM was trained on the binned hourly time series. d) The hidden state sequence was then generated for each validation participant using the trained HMM. e) The total dwell time was then calculated for each of these participants and compared to measures from questionnaires. f) Shows our main clinical findings: lower socially active dwell time in AD versus HCs, and an interaction between socially active dwell time and AD when predicting social functioning. z_t_: hidden state at time point, t, S1: State 1, S2: State 2, HC: healthy control, SZ: schizophrenia, AD: Alzheimer’s disease, SCC: Subjective cognitive complaints.

Where we use “one-hot” encoding for the latent variable, such that *Z*_*tn*_ = 1 if the latent variable at time *t* belongs to class *n*, and zero otherwise. The different components of this model are described in greater detail in the sections below.

The HMM model was implemented and fitted using the R package “depmixS4”.[22] During model training, the expectation-maximization algorithm is used to maximize the expected joint log-likelihood of the model parameters. The depmixS4 package allows for missing values in the dataset, which means that missing values are effectively omitted from the calculation of the log-likelihood, and also allows the specification of time-varying covariates that influence the transition probabilities as we outline below. Although depmixS4 also allows covariates to be specified over the starting probabilities, we do not explore this here. Each response variable (i.e. observed channel) was modeled using a multinomial distribution with an identity link function. As all of the input channels were binned into binary bins to manage the zero-inflation, this resulted in a binomial distribution for each response variable.

We investigated a range in the number of hidden states used by the HMM. As the input data included a total of five channels, a reasonable number of hidden states used by the HMM to achieve data compression ranged from two to four states. Due to this small range of number of hidden states, this hyperparameter was not formally optimized, but rather we select one main model for reporting and report results from the additional relevant models in Multimedia Appendix 2. We also report the Bayesian Information Criteria (BIC) for the various models. Additionally, we investigated the inclusion of the time of day (i.e. the hour) as a covariate in the model (i.e. over the transition probabilities), and used the BIC to determine whether to include this covariate in the models used for subsequent analyses. As the hour is recorded as ranging from zero (midnight) to 23 (11pm), the hour must be encoded so that it is not incorrectly implied that, for example, midnight is distant from 11pm. We therefore used one-hot encoding to encode the hour (where an indicator variable is used for each hour). We also investigated different seeds for model training, however this did not impact the likelihood of the model.

We then applied the trained HMM to the validation dataset and generated the hidden state sequences corresponding to these participants’ time series using the Viterbi algorithm. Note that this step did not involve retraining the model, and that the hidden state sequence is equal in length to the observed time series. For the alternative HMMs trained on the whole dataset, the hidden state sequences were generated for all participants, and subsequent investigations made for all participants.

### Hidden Markov Model Parameters and Measures

Various probabilities reflecting each of the hidden states are learned during model training, which can be used to describe the model and to understand what behaviors each of the hidden states are associated with. This includes emission, starting and transition probabilities:

#### Emission probability

The emission probability for each state refers to the probability that certain values in each of the observed channels are observed given that the sequence is in that hidden state, and can therefore be used to interpret what observed behaviors each hidden state represents. A state may give a high probability of observing activity in some observed behavioral channels and not others, and this can be identified with the emission probability. The emission probabilities of observed values **x**_*t*_ at time *t* given hidden states **z**_*t*_ are given by:

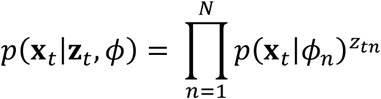

Where ϕ is a set of parameters governing the distribution of the observed data, N is the total number of hidden states in the model, i.e. in our case ranging from 2-4 for the different HMMs investigated.

#### Starting probability

The starting probability indicates the probability of beginning the sequence in each hidden state. If a time series often begins with the same observed values, then the hidden state corresponding to these values will have a high starting probability. The probability distribution giving the probability that each hidden state will be the first hidden state, **z**_1_is given by:

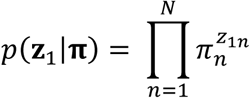

Where ℼ is the probability vector with elements π_*n*_ = *p*(*Z*_1*n*_ = 1).

#### Transition probability

The transition probability gives the probability of switching into another hidden state from each state (or the probability of staying in the same state). For example, for behaviors with long durations, the transition probability of staying in the associated hidden state may be high relative to the probability of transitioning to a non-related hidden state. The probability of transitioning into each hidden state at time *t* is dependent on the previous hidden state, and is given by:

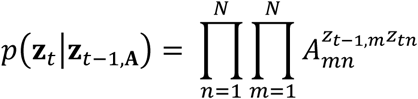

Where the elements of **A** are each of the transition probabilities such that *A*_*mn*_ = *p*(*Z*_*tn*_ = 1|*Z*_*t*−1,*m*_ = 1, *c*_*t*_) denotes the probability of transitioning from state *m* to state *n* at time *t* and we make it explicit that this can depend on a vector of time-varying covariates *c*_*t*_.

Additionally, other measures can be calculated from the hidden state sequence itself. In this study we focus on a measure referred to as the “dwell time”:

#### Dwell time

The dwell time per hidden state, also known as fractional occupancy, gives the percentage of time during which a state was occupied. This can be calculated for any desired level of granularity, for example, for all participants together, for each participant, for a specific time period, or for each instance a state is occupied. In this study we chose to calculate the total dwell time per participant, i.e. a single dwell time value per participant in the validation set reflecting the percentage of their time series that was spent in the socially active state. We chose this level of granularity as we had a single value from each social functioning/clinical measure available per participant, i.e. no repeated measures. As the validation set contained a range of data availability, any missing data timepoints were dropped from the time series after hidden state sequence generation, so that the calculation of dwell time only reflected the available data. As we focus on a 2-state model in this study, we concentrate solely on the total dwell time spent in one state (the “socially active” state), and do not refer to the dwell time of the other state in the analyses. For HMMs with more hidden states reported in Multimedia Appendix 2, we provide results for the states identified as socially active (see Figures S3, S5, S7, S9, S11 for the emission probabilities used to interpret each of these hidden states).

### Generalizability

As our principal model involves training on HCs, this could mean that the model is biased towards this population and not necessarily appropriate to use in other populations. To investigate whether a model trained on HCs can generalize sufficiently to the diagnostic groups we investigated two additional models: a model trained on all of the HCs, and a model trained on all of the remaining groups. We focused on two-state models here, and included the hour as a covariate over the transition probabilities following the same procedure as above. We compared the emission probabilities of these two models to establish whether equivalent hidden states were learnt, and then generated hidden state sequences for the participants in the diagnostic groups using both of these models. We then compared these hidden state sequences by evaluating the accuracy, sensitivity and specificity of the hidden state sequences provided by the HC model relative to the sequences provided by the diagnostic group model.

### Comparison of Total Dwell Time to Social and Clinical Measures

The total dwell times were used to predict two social measures using linear regression models: social functioning (SFS)[36] and loneliness[37] (available for participants in the PRISM study). For each of these measures, total dwell time, age, diagnostic group and interactions between diagnostic group and total dwell time were included as predictors. For SFS, separate models were also run for each of the diagnostic groups with age included as an additional predictor.

Total dwell times were then compared between the different diagnostic groups and HCs (available for participants in both PRISM and HO) using multinomial logistic regression, with total dwell time and age included as predictors. Sensitivity analyses of age were also carried out for each diagnostic group, due to the broad age range in HCs as a consequence of age-matching to both SZ and AD and expected possible generational differences in phone usage. For the SZ sensitivity analysis, the maximum SZ participant age was used as the maximum cut-off age for HCs (so age-matched HCs for the SZ age sensitivity analysis had a maximum age of 41), and for AD and SCC each respective minimum participant age was used as the minimum cut-off age for HCs (so age-matched HCs for the AD sensitivity analysis had a minimum age of 51, and for the SCC sensitivity analysis a minimum age of 44). Binomial logistic regression models were then run for each diagnostic group compared to their respective improved age-matched HCs.

Linear regression models were also run to predict cognitive impairment (MMSE) and schizophrenia symptoms (PANSS) (available for the AD (and HCs) and SZ participants in the PRISM study respectively) from total dwell times. For MMSE, total dwell time, age, diagnostic group and interactions between diagnostic group and total dwell time were included as predictors. In the case of PANSS scores, separate models were run to predict the total score and the subscores (positive, negative, general psychopathology and composite) from total dwell time and age.

To assist readability, we present the results from total dwell times from a single HMM in this paper. The equivalent results from additional HMMs can be found in Multimedia Appendix 2.

## Results

### Sample Statistics

This study utilized data from participants in the PRISM and HO datasets, which jointly contained 247 HCs, 18 participants with SZ, 26 with AD and 57 participants with SCC (see Table 1).

**Table 1:** Demographics of each of the diagnostic groups.

| Diagnostic group | Number | Age<br>(Mean<br>(SD)) | Gender | Dataset | Country | Education<br>years (Mean<br>(SD)) |
| --- | --- | --- | --- | --- | --- | --- |
| Healthy control | 247 | 59 (13) | F=140;<br>M=107 | PRISM=28;<br>HO=219 | NL=234;<br>ES=13 | 6 (4) |
| Schizophrenia | 18 | 31 (6) | F=7;<br>M=11 | PRISM=18;<br>HO=0 | NL=12;<br>ES=6 | 15 (3) |
| Alzheimer's<br>disease | 26 | 67 (7) | F=10;<br>M=16 | PRISM=19;<br>HO=7 | NL=18;<br>ES=8 | 13 (7) |
| Subjective<br>cognitive<br>complaints | 57 | 61 (7) | F=36;<br>M=21 | PRISM=0;<br>HO=57 | NL=57 | 5 (2) |

*SD: standard deviation; F: female, M: male; NL: the Netherlands, ES: Spain.*

Participants with AD and HCs were present in both datasets, whereas participants with SZ were provided by PRISM and participants with SCC were provided by HO.

In the PRISM and HO datasets, HCs were age matched to the diagnostic groups, with the PRISM sample being matched to both SZ and AD and the HO sample age-matched only to AD. After aggregation of datasets, this results in a bimodal age distribution. More specifically, due to the expected differences in age between participants with SZ and AD, the HCs are on average older than participants with SZ and younger than those with AD. However, note that the difference in age between the diagnostic groups is a consequence of aggregating multiple samples. From the age distributions presented in Figure 2, it is clear that the HC group spans the full range of each diagnostic group, and we also performed additional sensitivity analyses with HCs age-matched to the diagnostic groups to confirm group comparison findings. Training set and overall validation set age distributions are shown in Figure S8 in Multimedia Appendix 1.

**Figure 2:**
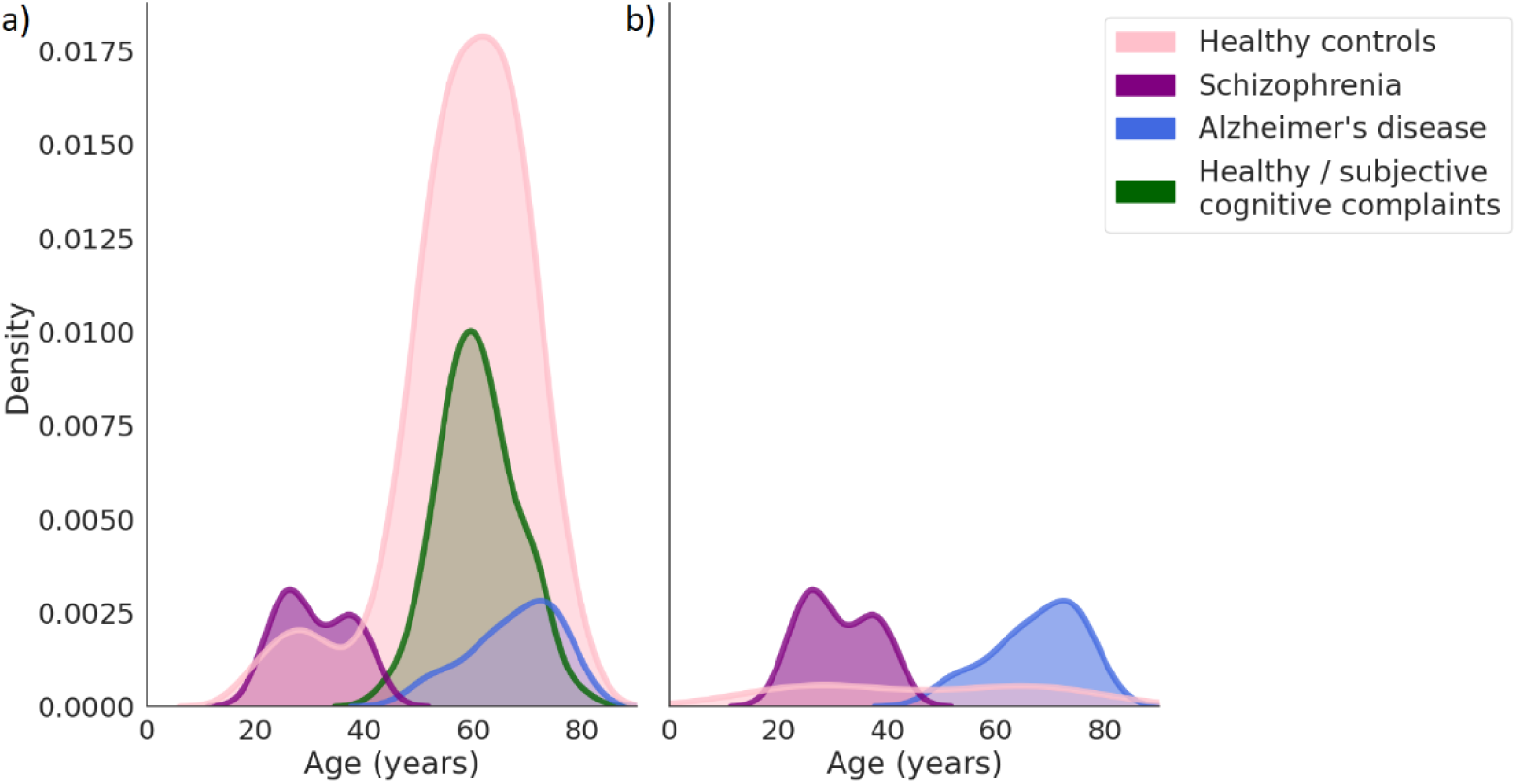
Age density distributions for validation participants. a) Distribution of ages for all validation participants. b) Distribution of ages for validation participants with social measures. Plotted using kernel density estimation.

PRISM data was collected across sites in the Netherlands and Spain, whilst HO data was collected solely in the Netherlands. PRISM recorded participant race, with nearly all participants identifying themselves as white, whereas HO did not report participant race. The demographics of the HCs, split by training versus validation set assignment, are provided in Table S1 in Multimedia Appendix 1.

### Hidden Markov Model Derivation and Interpretation

When training the HMM, the number of hidden states used by the model must be set. We evaluated 2-, 3– and 4-state models, which all converged. Generally as the number of hidden states increased the BIC improved, and it was also seen that including the hour as a covariate consistently improved the BIC (see Table S1 in Multimedia Appendix 2). We have chosen to primarily present results from a 2-state model for simplicity, but present equivalent results for other HMM variations in Multimedia Appendix 2. These alternative models varied in the number of hidden states (2-4) and the training set used (models trained on HCs with high data availability versus models trained on the entire dataset). For the models trained on all participants, total dwell times were also calculated for all participants (i.e. not only the validation set).

The emission probabilities of the states generated by the 2-state model are shown in Figure 3. Using these emission probabilities to interpret the hidden states, it is evident that they represent socially active and socially inactive states. That is, the second state (S2) corresponds to phone usage with a very high probability that communication apps are also being used by the participant. There is a smaller probability of social media usage and outgoing/incoming phone calls. Due to the use of communication methods in this state, such as calls and app usage, this hidden state is referred to as the “socially active” hidden state. The first state (S1) corresponds to a much smaller probability of phone usage, with the probability of all other channels near zero, and is referred to as the “socially inactive” hidden state. We show a demonstrative example of how the hidden states correspond to the observed channels in Figure 4, illustrating different observed channel configurations that can correspond to each of the hidden states.

**Figure 3:**
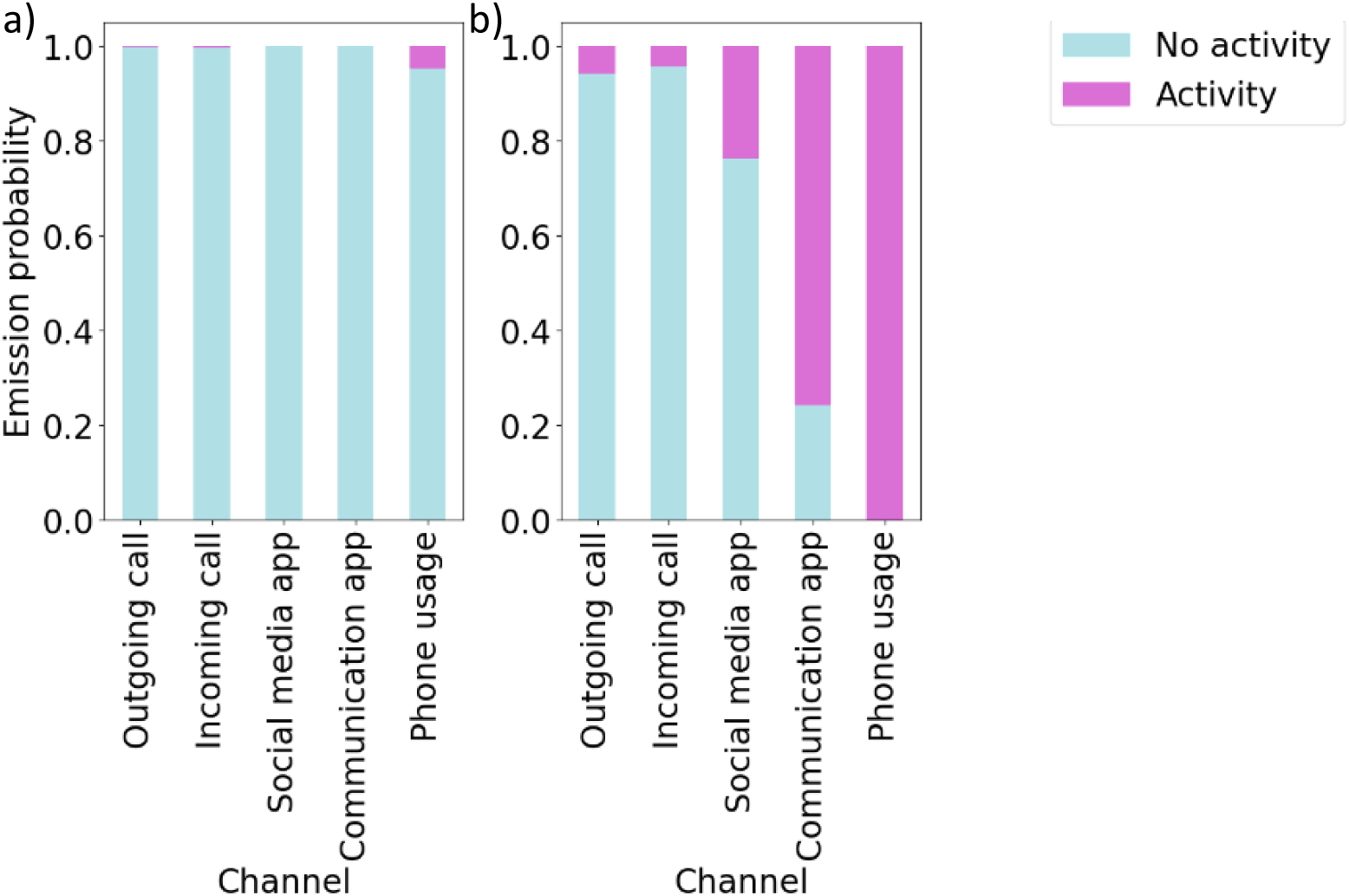
Emission probabilities of the selected 2-state model. Emission probabilities are provided for a) state 1 (S1) and b) state 2 (S2).

**Figure 4:**
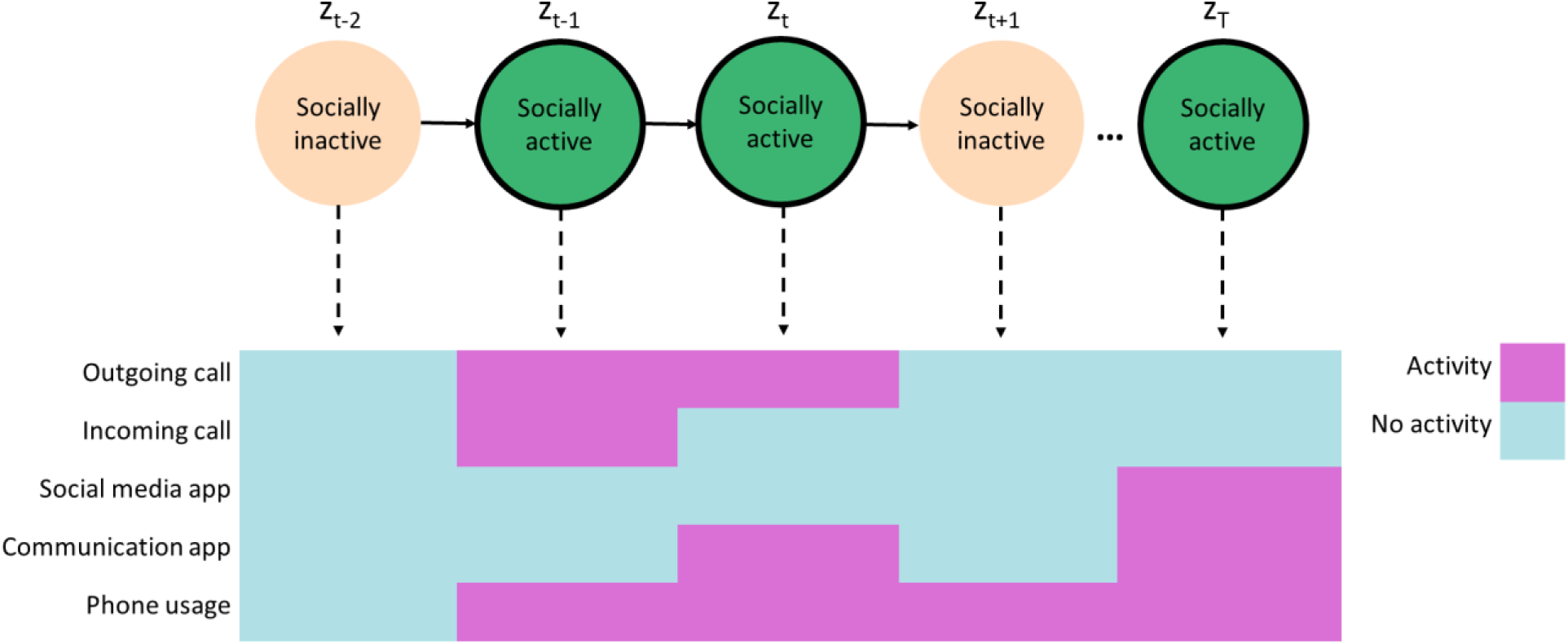
Examples of which behaviors may correspond to the hidden states. For the socially active state various social behaviors are displayed, including calls and app usage; in the socially inactive state there may be no phone usage, or phone usage without corresponding social behaviors.

After model training, the hidden state sequence corresponding to each participant’s time series was generated. The total dwell time for each validation participant can then be calculated from the hidden state sequence, with missing data in the validation set removed, and compared to clinical scores and diagnostic group. We chose to drop the missing portions from these time series after hidden state sequence generation due to the high rates of missingness for some participants. As the selected model only contains two states and the total dwell time (i.e. the proportion of time spent in each state) is a percentage value, only the dwell times corresponding to one of the states needed to be investigated. We therefore focus on the total dwell times from the “socially active” state, and so further reference to “total dwell time” derived from the HMM solely refers to dwell times in the socially active state.

An example of one participant’s hidden state sequence alongside the input sequence is shown in Figure 5, and an example of another participant can be seen in Figure 6. It is immediately apparent that the subject shown in Figure 5 spends considerably more time in the socially active state relative to the subject shown in Figure 6. It can be seen that the participants in both Figure 5 and Figure 6 oscillate quite frequently between the socially active and inactive states, which is not surprising due to expected diurnal variation (e.g. [39]). More clearly, higher social activity during the daytime and lower social activity during night-time can be seen in Figure 7, and in Figure 8 it can be seen that the probability of transitioning into the socially active state (state 2) from both the socially active and inactive states is increased during the daytime and drops off again in the evening. Additionally, the probability of starting a hidden state sequence in the socially active and inactive states were 0.26 and 0.74 respectively, showing that it is more probable to begin the time series in the socially inactive state. This is to be expected as all of the time series began at midnight, so many participants would have been asleep.

**Figure 5:**
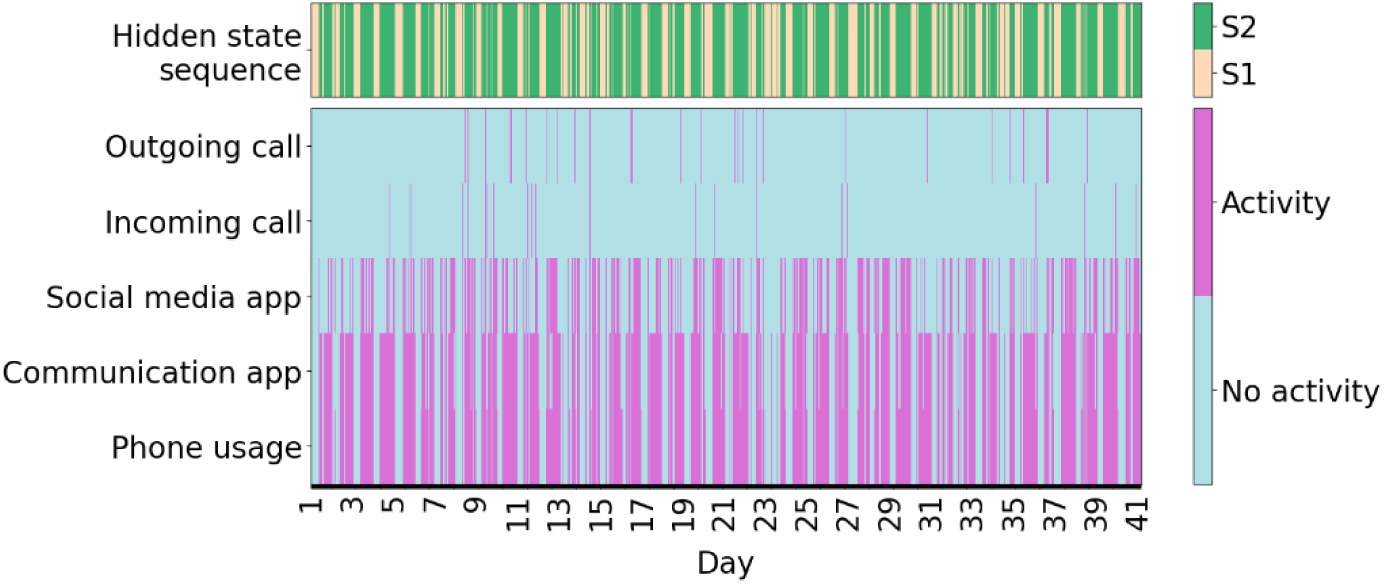
Example time series with high social activity. The observed time series composed of hourly bins (bottom five rows) of a participant compared with their corresponding predicted hidden state sequence (top row). S1: State 1 (socially inactive state), S2: State 2 (socially active state).

**Figure 6:**
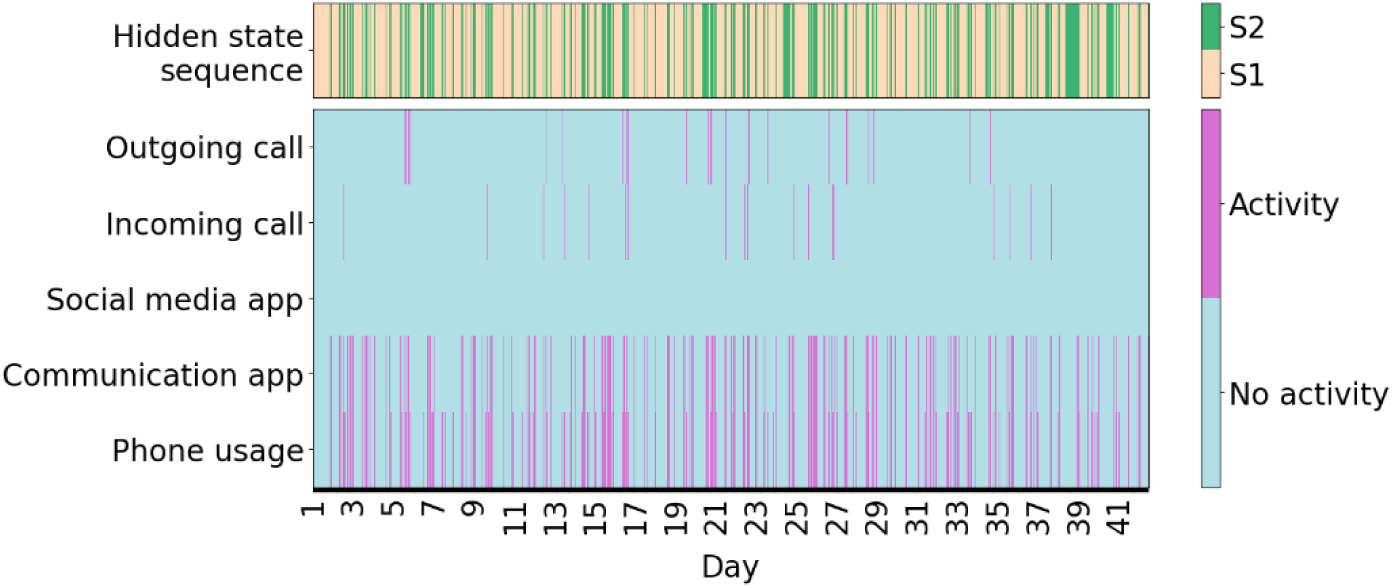
Example time series with low social activity. The observed time series composed of hourly bins (bottom five rows) of another participant compared with their corresponding predicted hidden state sequence (top row). S1: State 1 (socially inactive state), S2: State 2 (socially active state).

**Figure 7:**
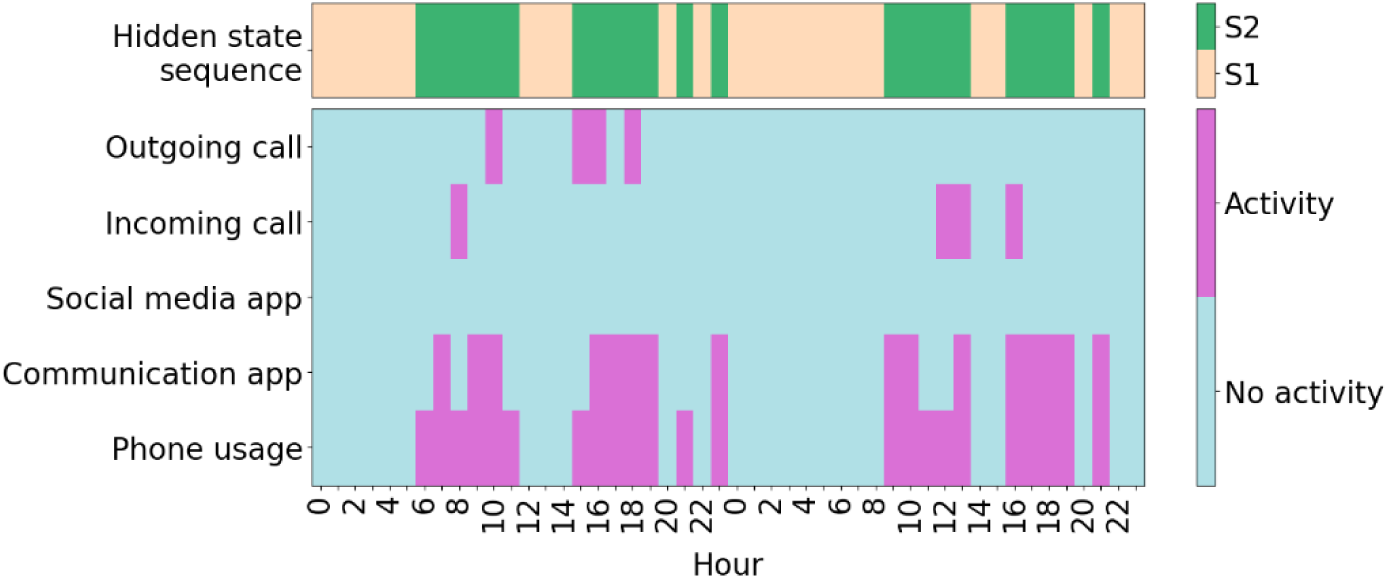
An example of a 2-day period of a participant’s time series. This participant showed higher social activity during the daytime than the nighttime. S1: State 1 (socially inactive state), S2: State 2 (socially active state), 0: midnight.

**Figure 8:**
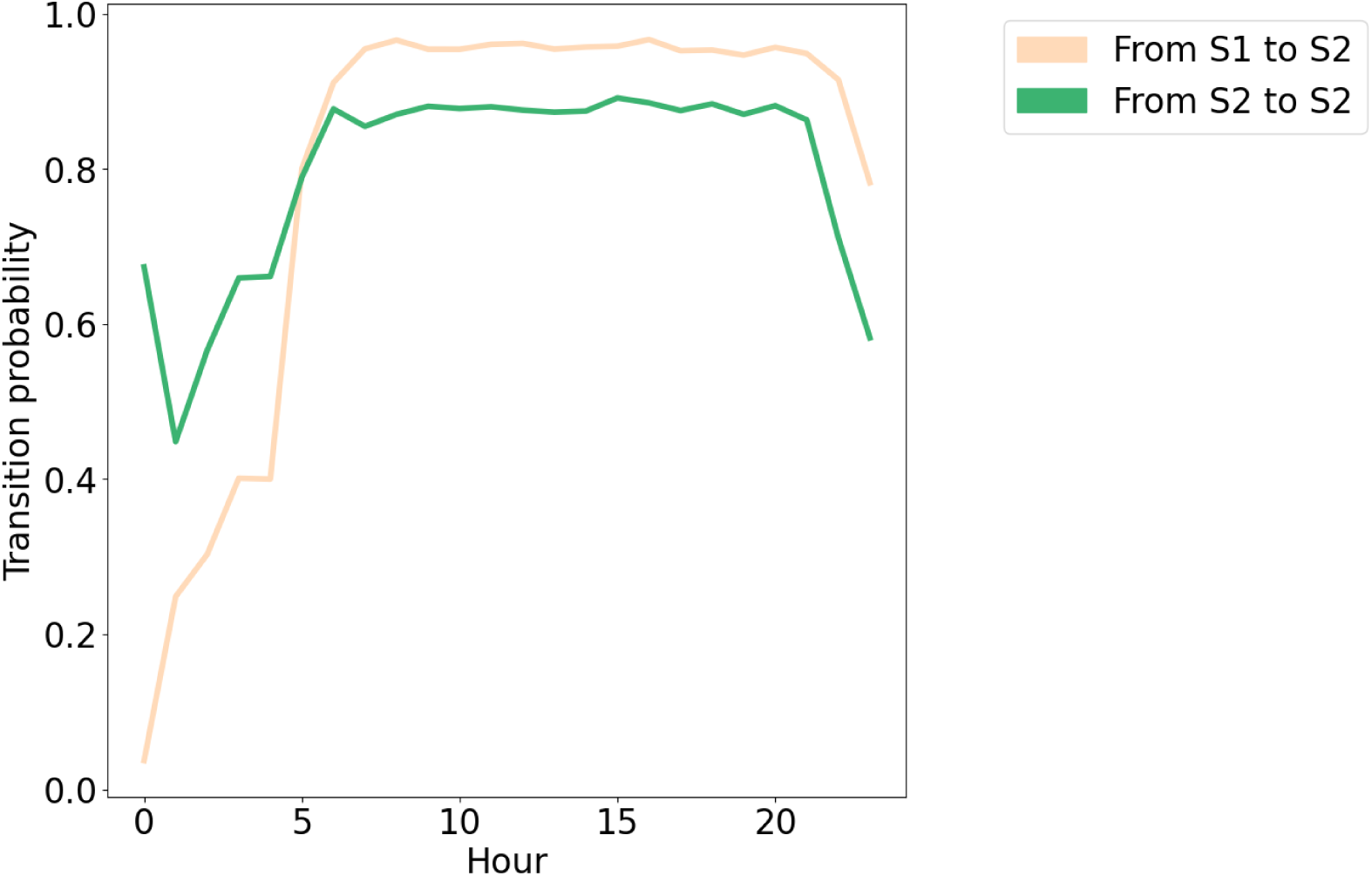
The probability of transitioning into the socially active state from each state, for each hour in the day. S1: State 1 (socially inactive state), S2: State 2 (socially active state), 0: midnight.

### Generalizability

To investigate the generalizability of the approach of training the HMM using HCs and evaluating in other diagnostic groups, we compared the hidden state sequences of participants in the diagnostic groups generated from two different models: a model trained solely on these participants, and a model trained solely on HCs. We found that both of these models produced very similar hidden states (see Figures S1 and S2 in Multimedia Appendix 2), with state 1 in each model corresponding to social activity. We therefore did not need to relabel the hidden states before comparing the models. More specifically, considering the hidden state sequences from the model trained on all diagnostic groups as the “true” sequence, we found that the hidden state sequences from the model trained on HCs had an overall accuracy of 0.91, a sensitivity of being in the socially active state of 1.0 and specificity of 0.86. Overall this suggests that an HMM trained on HCs can generalize adequately to the diagnostic groups in this analysis.

### Measures of Social Functioning and Loneliness

For validation purposes, we make use of a measure of social functioning for each participant in the PRISM dataset, namely the Social Functioning Scale (SFS)[36] (see Figures S9 and S10 in Multimedia Appendix 1 for score distributions). We therefore investigated possible relationships between social functioning and total socially active dwell times for participants with SFS scores available. The number of participants in each group is small, so we consider our results to be preliminary indicators of possible relationships between the HMM-derived digital phenotypes and social functioning.

To investigate the relationship between social functioning and total dwell time, we ran linear regression models that predicted SFS score from total dwell time, age, diagnostic group and interactions between diagnostic group and total dwell time. HCs were taken as the reference group. FDR corrected *P* values (considering six tests) are presented with results considered significant at *P*<.05 (see Table 2). A significant interaction between AD and total dwell time was identified (FDR corrected *P* value=0.02) (see Figure 9), however no significant main effect of total dwell time was found. This result was robust across HMMs with different numbers of states and regardless of whether the model was trained on high data availability HCs and assessed on withheld participants, or trained and evaluated for all participants (see Tables S2-S7 in Multimedia Appendix 2). Additionally, a significant main effect of SZ group relative to HCs was seen (FDR corrected *P* value=0.02), with lower SFS scores seen in SZ, but no significant main effect of AD was seen.

**Figure 9:**
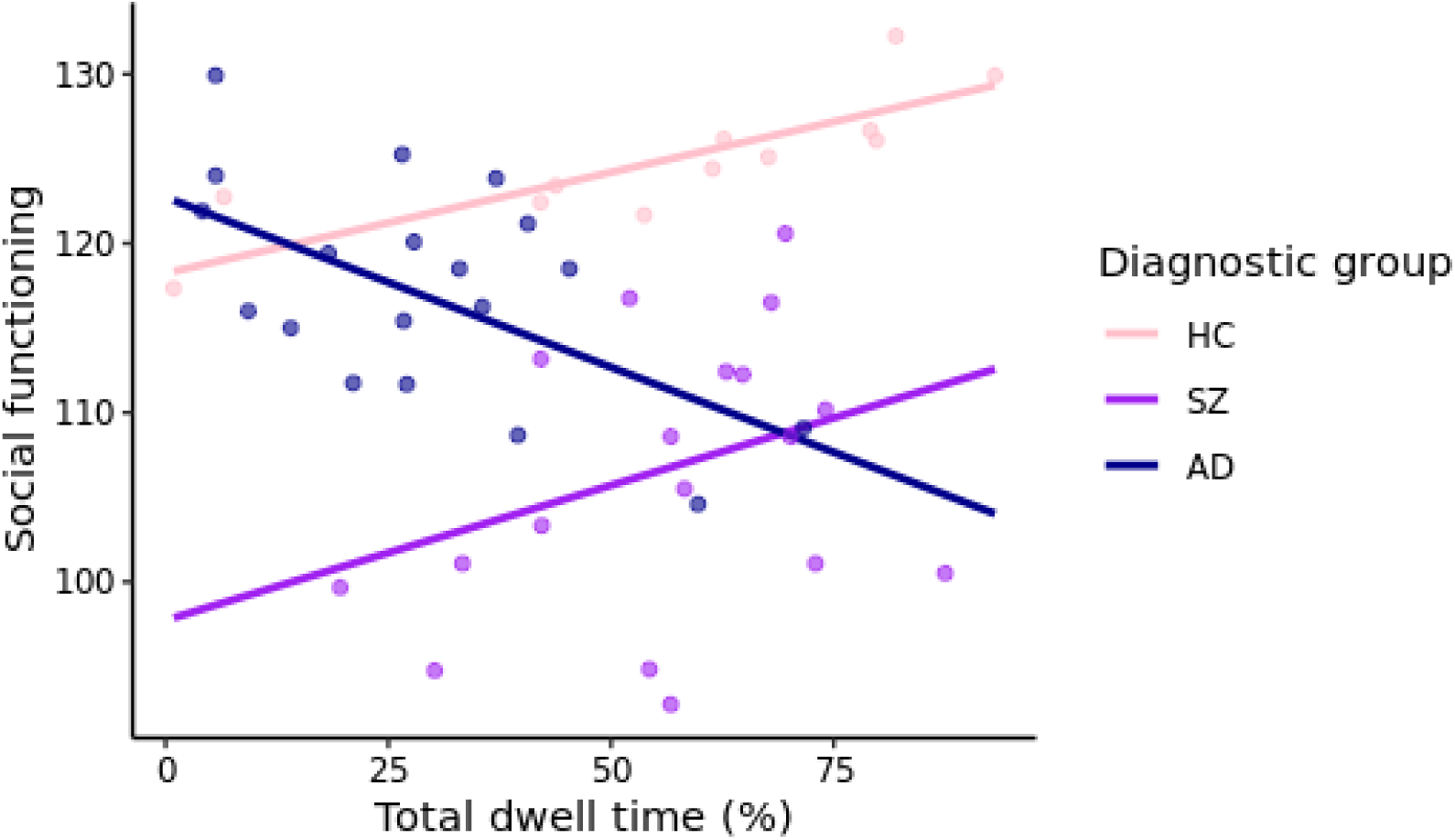
Social functioning scale score against total dwell time, with interactions displayed for the different groups (HC: healthy controls, SZ: schizophrenia, AD: Alzheimer’s disease).

**Table 2:** Results from a linear regression model predicting SFS score from total dwell time, age and group, where healthy controls (n=12) were the reference group.

| Predictor | Coefficient | Standard error | *t* value | *P* value | FDR corrected *P* value |
| --- | --- | --- | --- | --- | --- |
| Age | 0.0269 | 0.0788 | 0.3406 | .74 | 1.0 |
| Group: Schizophrenia (n=18) | -20.5052 | 6.6684 | -3.0750 | .004 | .02 |
| Group: Alzheimer's disease (n=19) | 4.4857 | 4.8877 | 0.9178 | .36 | 1.0 |
| Total dwell time | 0.1193 | 0.0663 | 1.7982 | .08 | .48 |
| Interaction: Schizophrenia and total dwell time | 0.0401 | 0.1069 | 0.3757 | .71 | 1.0 |
| Interaction: Alzheimer's disease and total dwell time | -0.3201 | 0.1020 | -3.1384 | .003 | .02 |

Linear regression models were also run within the different diagnostic groups to investigate possible within-group relationships between SFS scores and total dwell times. Separate models were run for each of the diagnostic groups in the validation set, with age included as an additional predictor in the models. FDR corrected *P*-values (considering three tests) are presented in Table S2 in Multimedia Appendix 1, with results considered significant at *P*<.05. A significant positive relationship between social functioning and dwell times was found for the HCs (FDR corrected *P* value=.005), with every one percent increase in total dwell time corresponding to a 0.1153 increase in SFS score, however this relationship was not seen when evaluating the entire HC group (using the HMM trained on all participants) and is expected to be due to sampling variation rather than differences in the learnt HMM parameters. No significant relationship was found for the other diagnostic groups.

A measure of loneliness[37] was also provided for the PRISM participants, however no significant relationship between loneliness and total dwell times was found. The results from this linear regression model are presented in Table S3 in Multimedia Appendix 1, as well as histograms of the distribution of loneliness scores (Figures S11 and S12).

### Diagnostic Group

A multinomial logistic regression model was run to investigate differences in total socially active dwell time between the different diagnostic groups and the HC group in the validation set (i.e. the reference category) (see Figure 10). Age was again included as an additional predictor in the model (age-related results are presented in Table S4 in Multimedia Appendix 1), and FDR corrected *P*-values (considering three tests) are presented to provide an indicator of significance at *P*<.05 (Table 3). Total dwell time was found to be a significant predictor of AD relative to HCs (FDR corrected *P* value<.001); participants with AD generally showed lower dwell times (i.e. spending less time in the socially active state) relative to HCs (odds ratio=0.9483). This relationship was also seen across almost all equivalent socially active hidden states of the additional HMMs considered, with results of these models presented in Tables S8, S10, S12, S14, S16, S18 in Multimedia Appendix 2. For the SCC group, lower total dwell times were also observed relative to HCs (FDR corrected *P* value=0.004, odds ratio= 0.9742). However, this result was less robust when considering the other HMM variations. No significant relationship of total dwell time on SZ group was found relative to HCs. Due to the broad age range of HCs, sensitivity analyses of age were carried out for each diagnostic group (Table S5 in Multimedia Appendix 1), with a subsample of HCs age-matched to each respective diagnostic group, with the AD result remaining significant (FDR corrected *P* value<.001), as well as the SCC result (FDR corrected *P* value=.003) (see Tables S9, S11, S13, S15, S17, S19 in Multimedia Appendix 2 for all sensitivity analyses).

**Figure 10:**
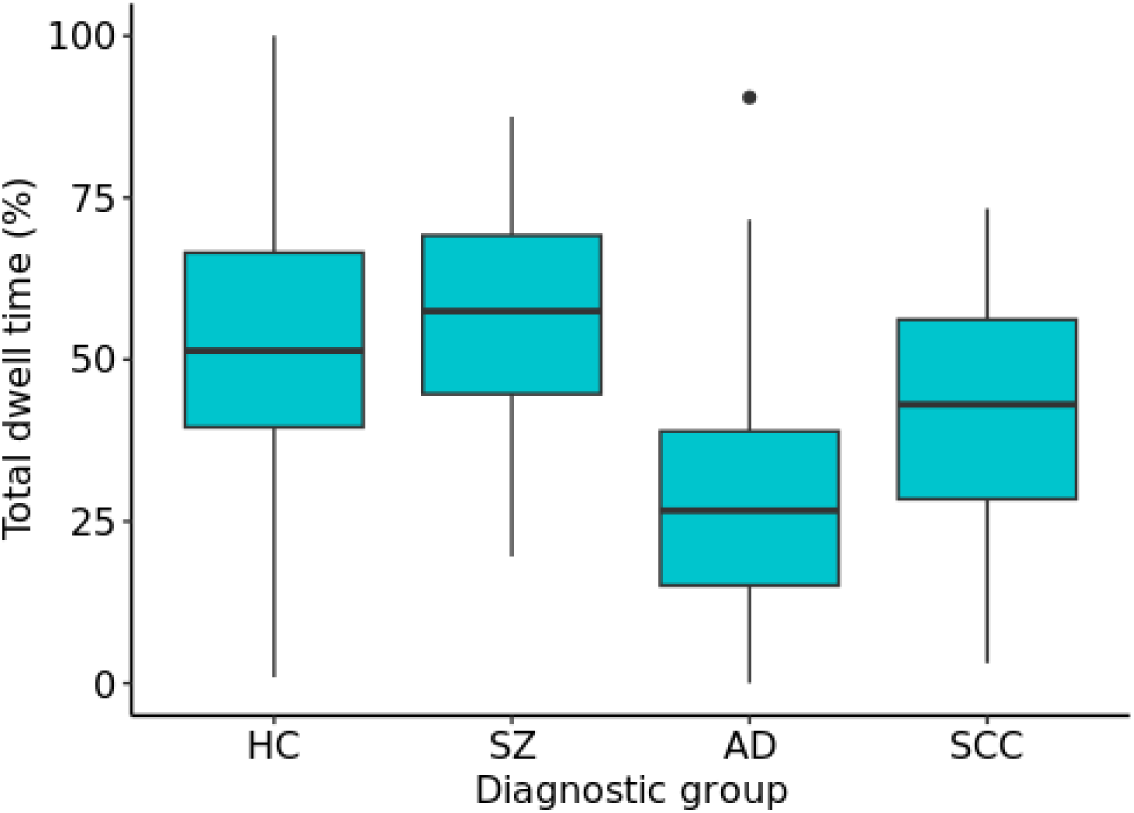
A box plot of the total dwell times per participant for the different diagnostic groups. There is a significant difference between the HC and AD groups, and the HC and SCC groups. HC: healthy control, SZ: schizophrenia, AD: Alzheimer’s disease, SCC: Subjective cognitive complaints.

**Table 3:** Results from a multinomial logistic regression model predicting diagnostic group (versus healthy controls (n=156)) using total dwell time. Age was also included as a predictor.

| Group | Coefficient | Standard error | Odds | z value | P value | FDR corrected P value |
| --- | --- | --- | --- | --- | --- | --- |
| Schizophrenia (n=18) | -0.0133 | 0.0172 | 0.9867 | -0.7763 | .44 | 1.0 |
| Alzheimer's disease (n=26) | -0.0531 | 0.0132 | 0.9483 | -4.0097 | <.001 | <.001 |
| Subjective cognitive complaints (n=57) | -0.0262 | 0.0082 | 0.9742 | -3.1846 | .001 | .004 |

### Further Clinical Measures

For participants with AD and the HCs in the PRISM dataset, Mini-Mental State Examination (MMSE)[35] scores, measuring cognitive impairment, were provided. No significant effect of total dwell time or age, nor a significant interaction between dwell time and diagnostic group was found, although there was a significant effect of diagnostic group (see Table S6 in Multimedia Appendix 1, as well as score distributions in Figures S13 and S14 in Multimedia Appendix 1). This was expected given the inclusion criteria of the study.

The PRISM dataset also provided Positive and Negative Syndrome Scale (PANSS)[34] scores for participants with SZ, however no significant relationships between any of the PANSS scores (positive, negative, general psychopathology, composite and total) and total dwell time were found. The results from these linear regression models are presented in Table S7 in Multimedia Appendix 1, as well as histograms of the distribution of PANSS scores per subscale (Figure S15).

## Discussion

### Principal Results

A central aim of digital phenotyping is to develop objective measures that can be used to monitor clinically-relevant behaviors and symptom changes. In this study we proposed a method for deriving meaningful, interpretable digital phenotypes using the HMM, a time series model that can accommodate missingness. We applied this model to general phone usage and communication smartphone measures, calculating the total socially active dwell time phenotyped by the HMM. Our smartphone measures were collected passively, reducing burden on participants, and we protected participant privacy by abstracting app measures to descriptive levels, without collecting content. We investigated the association of the total socially active dwell time with various social and clinical measures, including diagnostic group and a questionnaire on social functioning (SFS). We found that 2-4 state HMMs provided comparable socially active states, which showed consistent results when investigating the relationships between the total dwell time and the social and clinical measures. We observed a significant difference in the HMM-derived total “socially active” dwell times between HCs and participants with AD, with participants with AD exhibiting lower total dwell times. This difference was robust to age sensitivity analysis and across different HMM variations (in terms of the number of hidden states and the training set used). A significant interaction between total dwell times and AD label was also observed for social functioning.

The HMM has several strengths: it uses lower-dimensional hidden states to represent the various observed behaviors, which can be easily interpreted for each state using the emission probabilities (Figure 3). The socially active state could be interpreted as being linked to observed communication-related behaviors, whilst the socially inactive state reflected a lack of these behaviors, such as other kinds of, or no, phone usage. Transitions between these hidden states were indicative of behavioral changes throughout time, for example daily behavioral patterns (Figure 7). It was seen that during the daytime it was more likely for participants to transition to the socially active state than during the nighttime (Figure 8). Hidden states may allow for some individual behaviors to be represented as comparable behaviors. For example, Figure 6 shows a time series with no social media usage, whereas Figure 5 shows highly recurrent social media usage, and both of these participants can have their respective behaviors represented using the socially active state despite individual differences in what social activity may mean for each participant. This type of modeling approach can therefore allow for a certain amount of flexibility in the behaviors of the participants, dependent on the number of hidden states used in the model.

A summary measure of the HMM, the total socially active dwell time, was calculated per validation participant so that a model-derived digital phenotype could be compared to clinical and social measures. The observed difference in total dwell time between participants with AD and HCs, with AD dwell times lower than HCs, is consistent with the understanding that AD is associated with impaired social functioning,[1] and demonstrates a potential objective measure of this difference. A significant interaction between total dwell time and AD was also seen when predicting social functioning, further demonstrating this.

Similarly to the AD group, differences in dwell time relative to HCs were also observed for SCC participants, however these differences were less robust across HMM variations. Differences in dwell time were not observed for participants with SZ. These results may be unsurprising as by definition SCC participants are very similar to HCs, with the difference in inclusion criteria being that SCC participants experience memory complaints. Similarly, the participants with SZ did, for the most part, exhibit quite low symptom severity. The number of participants with SZ was also small. Whilst the PRISM study only placed exclusion criteria on positive symptoms (to exclude psychosis), the negative symptoms in the sample did not turn out to be very severe either, and overall most participants could be classified as “mildly ill” based on their total PANSS score.[40] This is indicative of a selection of less affected patients. The mild PANSS scores, as well as low loneliness scores, may also contribute to the absence of an identified relationship between these scales and total dwell time. When investigating social functioning within the different groups, significant relationships between social functioning and total dwell time for participants with AD and SZ were also not observed. It is possible that participants with AD and SZ may overestimate their social functioning,[41] which could be reflected in their self-report SFS scores. This may complicate any possible relationship between this social functioning measure and dwell time for these groups. A further interesting factor that could affect these relationships is the impact of different symptom profiles on dwell time.

### Future Directions

To expand upon the current work, the HMM method could be applied in a larger population of SZ participants exhibiting broader symptom severity and different symptom profiles. Given the reluctance of many people with acute psychotic symptoms to being monitored, it may be necessary to monitor participants for a longer period of time, beginning with low symptom severity at study enrolment, to allow for more fluctuations in symptom severity to be observed (e.g. [20]). The HMM method can also be applied to other disorders, including Major Depressive Disorder (to be included in PRISM 2). A wider range of smartphone channels can also be included in the HMM, for example calls could be encoded to reflect the variation in who is called/is calling each hour. With a larger number of input channels, the derived hidden states could reflect more specific behavioral states. The optimal number of hidden states may then be driven both by the number of input channels, and the underlying behavioral states of the participants themselves. With a higher order model, the hidden states’ emission probabilities would not necessarily correspond to distinct single behaviors; for example, with the inclusion of GPS channels there could be two hidden states that correspond to time spent at home, with one state also reflecting communication activities and the other reflecting no communication.

In our analysis, each hidden state sequence was generated per participant, but total dwell time comparisons were only made between groups. To shift towards individual predictions (for example predicting symptom scores or relapse along the time series), the dwell time for windows of the sequence, or potentially the sequence likelihood, could be extracted and changes along the time series evaluated. This would also maintain the time component of the analysis; our current analysis uses a time series model but then compares a summary HMM measure to clinical measures. For clinical applications, the eventual goal would be to be able to make individual predictions along the time series. For this goal it may be beneficial to include group-specific transition probabilities. It could also be beneficial to allow some individual parameters, for example individualized transition probabilities could be considered. The choice of individual versus group-specific parameters may depend on the data available per individual. Zero-inflated distributions for the various channels could also be investigated as an alternative to binning the data into activity versus no activity bins.

Using our HMM trained on high data availability HCs as a reference group, we could identify differences in total socially active dwell time between the HCs and AD, as well as between HCs and SCCs, and an interaction between AD group and total dwell time on social functioning scores. These findings were also observed in models trained using the entire dataset. However by maintaining our dataset split in future analyses, this would allow us to look into the likelihood as a tool for identifying time series for which the model is a “poor fit”, i.e. deviates from the reference group. This could be useful in datasets where, for example, the aim is to identify relapse, but for which there are not necessarily many examples of the relapse periods available that can be utilized in model training. These deviations could potentially be used to identify anomalous time series.

To improve the management of missing data, there are several more avenues that can be explored. Data is often expected to be missing due to technical difficulties, but it is also possible that data can be missing due to user behavior, for example if the user switches the phone off, turns on flight mode or deletes the app from their phone. It is also possible that a phone running out of battery could be correlated with certain activities carried out by the user and/or with certain times of the day, meaning the missingness is caused indirectly by user behavior. Future studies could consider recording the direct behaviors (which would currently be more feasible with Android phones, rather than iOS), to provide a better indicator of data missing due to technical difficulties versus user behavior.

### Limitations

Due to high rates of missingness, we made four main decisions to handle missing data: (1) to focus model training on high data availability time series, (2) to use a model that can accommodate missing data, (3) to exclude GPS channels from this time series analysis due to low levels of data availability affecting these specific channels and (4) to exclude missing timepoints from the validation time series after hidden state sequence generation. Whilst we view decisions (1) and (2) as useful strategies for managing missing data, decision (3), and to a lesser degree decision (4), were unfortunate consequences that in future studies should be avoided with improved data collection. The datasets used in this study were collected with early versions of Behapp, and throughout data collection no indicator of missingness was known. Indicators of missing data were developed retrospectively using WiFi and GPS sampling frequencies to assist analyses of these time series. Incoming data monitoring has now been improved in more recent Behapp versions, as well as the overall data collection process. Researchers using Behapp can therefore now track data collection as it is ongoing, and take action if sustained periods of data are missing. This could involve contacting participants to ensure they have not accidentally disabled desired functionalities for sustained periods. Additionally, whilst the analysis package we used assumes that data is missing at random, and therefore equally likely to be missing across the different hidden states, it is possible that this is not the case and that missingness may vary across hidden states. For example, in cases where missingness may be due to user behavior affecting battery consumption.

For interpretation purposes, we have named the two hidden states as “socially active” and “socially inactive”. However, a person could, of course, be socially active offline without using their phone. For example, a person may be socializing with friends at home without using their phone. We therefore acknowledge limits to our naming convention, and recommend caution when interpreting hidden states. Other sensors could be used to give an indicator of other people in the participant’s vicinity, such as Bluetooth,[42] but passive smartphone data will nevertheless remain somewhat of a proxy for social activity. In a similar vein, we used the App Store classification to group apps, but participants may use the apps for purposes other than this classification (e.g. some people use Instagram for communication, and less so for social media). Whilst in our 2-state model these discrepancies would be inconsequential, with a larger number of hidden states these discrepancies could potentially lead to misleading interpretations of a person’s behavior. In a clinical setting, the patient’s behaviors could be discussed with the clinician at the beginning of Behapp usage to assist in understanding and interpreting their personal digital phenotypes.

Additionally, it is worth noting that by using a reference class approach we do restrict the model to only learning hidden states present in the reference group (as well as transition and starting probabilities associated with the reference). Whilst we also trained the HMM on all participants (see Multimedia Appendix 2) and found that the hidden states present in our HMM trained on high data availability HCs were highly comparable to the hidden states in HMMs trained on the whole dataset, for other datasets (such as those with a larger number of input channels) it may be that those in a clinical group could exhibit different hidden states, or that if a clinical group in remission or with low symptom severity is used for model training that states associated with relapse or high symptom severity would not be learned by the model. Therefore, the dataset used for training the model and the subsequent analysis steps must be considered, as this restricts the hidden states that are learned by the model. In such cases accepting that the trained model may not be a “good fit” for the withheld data could be something that could be used to help rather than hinder the analysis, by looking for deviations in the likelihood of such data with respect to the learned HMM.

## Conclusions

Smartphone-based digital phenotyping is a promising tool for monitoring and predicting mental health outcomes. However, methods are needed for managing this multi-faceted time series smartphone data. We proposed the use of an HMM to model digital phenotyping time series, as this method can (1) combine different behavioral features, (2) reflect temporal behavioral changes (3) be easily interpreted and (4) manage missingness. We developed a 2-state model that represented various smartphone channels as “socially active” and “socially inactive” states, and calculated the total socially active dwell time for each participant’s time series. We identified a significant difference between HC and AD dwell times, with AD dwell times lower than HCs, showing how this HMM-derived digital phenotype may be a useful measure to indicate differences in social functioning. We also observed a significant interaction between total dwell time and AD group when predicting social functioning. The HMM is an interpretable method to model behavior based on digital phenotyping data and with further development provides an appealing approach for making clinical predictions of symptom changes and relapse across a range of neuropsychiatric diseases.

## Supporting information

Multimedia Appendix 1. Supplementary Materials

Multimedia Appendix 2. Supplementary Materials

## Data Availability

The datasets analysed during the current study are available from the corresponding author on
reasonable request.

## Acknowledgements

This study was funded by the European Research Council (consolidator grant 101001118). The Dutch Brain Research Registry (Hersenonderzoek.nl) is supported by ZonMw-Memorabel (project no 73305095003), Alzheimer Nederland, Amsterdam Neuroscience, and Hersenstichting (Dutch Brain Foundation). The PRISM project has received funding from the Innovative Medicines Initiative 2 Joint Undertaking under grant agreement 115916. This Joint Undertaking receives support from the European Union’s Horizon 2020 research and innovation programme and EFPIA. This study reflects only the authors’ view and the European Commission is not responsible for any use that may be made of the information it contains.

## Authors’ Contributions

IEL: conceptualization, formal analysis, methodology, software, visualization, writing – original draft

AC: data curation, investigation, software, writing – review & editing RJ: data curation, software, writing – review & editing

LMR: data curation, writing – review & editing PJV: data curation, writing – review & editing

MJHK: conceptualization, funding acquisition, project administration, writing – review & editing

CB: conceptualization, supervision, writing – review & editing HGR: conceptualization, supervision, writing – review & editing

AFM: conceptualization, funding acquisition, methodology, supervision, writing – review & editing

## Multimedia Appendix 1

Supporting results for the main HMM variation that is reported, as well as histograms reflecting different aspects of the Behapp data and participant scores.

## Multimedia Appendix 2

Results from additional HMM variations, including HMMs trained on the entire dataset and HMMs with 3-4 hidden states.

## Abbreviations

AD: Alzheimer’s disease
DSM-IV: Diagnostic and Statistical Manual of Mental Disorders
HC: Healthy Control
HMM: Hidden Markov Model
HO: Hersenonderzoek
MDD: Major Depressive Disorder
MINI: Mini-International Neuropsychiatric Interview
MMSE: Mini-Mental State Examination
NIAAA: National Institute on Aging and the Alzheimer’s Association
S1: State 1 (socially active state)
S2: State 2 (socially inactive state)
SCC: Subjective cognitive complaints
SFS: Social Functioning Scale
SZ: Schizophrenia
PANSS: Positive and Negative Syndrome Scale
PRISM: Psychiatric Ratings using Intermediate Stratified Markers

