## Supplementary material for "Uncovering Social States in Healthy and Clinical Populations Using Digital Phenotyping and Hidden Markov Models: Observational Study": Multimedia Appendix 1. Supplementary Materials

#### Tables

The results in these tables relate to the main HMM reported in the manuscript, i.e. the two-state model trained on withheld healthy controls with high data availability and assessed on the validation set. For additional tables of results from alternative HMM variations (i.e. different number of hidden states, trained on whole dataset) see Multimedia Appendix 2.

*Table S1: Demographics of healthy controls with training/validation split.*

| Set | Number | Age (Mean (SD)) | Gender | Dataset | Country | Education years (Mean (SD)) |
|---|---|---|---|---|---|---|
| Training | 91 | 60 (14) | F=47; M=44 | PRISM=16; HO=75 | NL=83; ES=8 | 7 (5) |
| Validation | 156 | 58 (12) | F=93; M=63 | PRISM=12; HO=144 | NL=151; ES=5 | 6 (3) |

**SD: standard deviation; F: female, M: male; NL: the Netherlands, ES: Spain.*

*Table S2: Results from linear regression models predicting SFS score from total dwell time and age, with separate models run for the different groups.*

| Group | Predictor | Coefficient | Standard error | *t* value | *P* value | FDR corrected *P* value |
|---|---|---|---|---|---|---|
| Healthy controls (n=12) | Total dwell time | 0.1153 | 0.0257 | 4.4784 | .002 | .005 |
| | Age | 0.0100 | 0.0357 | 0.2810 | .79 | 1.0 |
| Schizophrenia (n=18) | Total dwell time | 0.1657 | 0.1166 | 1.4211 | .18 | .53 |
| | Age | 0.1164 | 0.3188 | 0.3651 | .72 | 1.0 |
| Alzheimer's disease (n=19) | Total dwell time | -0.1994 | 0.0752 | -2.6501 | .02 | .052 |
| | Age | 0.0380 | 0.1696 | 0.2239 | .83 | 1.0 |

Note that the significant result observed for healthy controls here was not observed when considering all healthy controls using the HMMs trained on all participants.

*Table S3: Results from a linear regression model predicting Loneliness score from total dwell time, age and group where healthy controls (n=12) were the reference group.*

| Predictor | Coefficient | Standard error | *t* value | *P* value |
|---|---|---|---|---|
| Age | 0.0052 | 0.0034 | 1.5480 | .13 |
| Group: Schizophrenia (n=18) | 0.4452 | 0.2859 | 1.5573 | .13 |
| Group: Alzheimer's disease (n=19) | -0.2691 | 0.2096 | -1.2841 | .21 |
| Total dwell time | -0.0016 | 0.0028 | -0.5668 | .57 |
| Interaction: Schizophrenia and total dwell time | -0.0004 | 0.0046 | -0.0909 | .93 |
| Interaction: Alzheimer's disease and total dwell time | 0.0031 | 0.0044 | 0.7200 | .48 |

The other HMMs provided comparable results.

*Table S4: Results from the age predictor in the multinomial logistic regression model predicting diagnostic group (versus healthy controls (n=156)) from total dwell time and age.*

| Group | Coefficient | Standard error | Odds | *z* value | *P* value | FDR corrected *P* value |
|---|---|---|---|---|---|---|
| Schizophrenia (n=18) | -0.1660 | 0.0298 | 0.8470 | -5.5737 | <.001 | <.001 |
| Alzheimer's disease (n=26) | 0.0801 | 0.0295 | 1.0834 | 2.7158 | .007 | .02 |
| Subjective cognitive complaints (n=57) | 0.0125 | 0.0163 | 1.0126 | 0.7665 | .44 | 1.0 |

*Table S5: Binomial logistic regression age sensitivity analysis results. A model was run per diagnostic group, where the reference group for each model was age-matched healthy controls. Age cut-offs for healthy controls (HC) matched to each diagnostic group (schizophrenia (SZ), Alzheimer's disease (AD), subjective cognitive complaints (SCC)): HC vs SZ, Age < 42 (HC n=12); HC vs AD, Age > 50 (HC n=130); HC vs SCC, Age > 43 (HC n=142).*

| Group | Predictor | Coefficient | Standard error | Odds | *z* value | *P* value | FDR corrected *P* value |
|---|---|---|---|---|---|---|---|
| Schizophrenia (n=18) | Total dwell time | -0.0036 | 0.0190 | 0.9964 | -0.1882 | .85 | 1.0 |

| | | | | | | | |
|---|---|---|---|---|---|---|---|
| | Age | 0.0621 | 0.0633 | 1.0641 | 0.9810 | .33 | 0.98 |
| Alzheimer’s disease (n=26) | Total dwell time | -0.0521 | 0.0136 | 0.9493 | -3.8421 | <.001 | <.001 |
| | Age | 0.0527 | 0.0327 | 1.0541 | 1.6134 | .11 | .32 |
| Subjective cognitive complaints (n=57) | Total dwell time | -0.0283 | 0.0086 | 0.9721 | -3.2932 | <.001 | .003 |
| | Age | -0.0199 | 0.0210 | 0.9803 | -0.9436 | .35 | 1.0 |

*Table S6: Results from a linear regression model predicting MMSE score from total dwell time, age and group where healthy controls (n=12) were the reference group. P values were corrected considering four tests.*

| Predictor | Coefficient | Standard error | *t* value | *P* value | FDR corrected *P* value |
|---|---|---|---|---|---|
| Age | -0.0045 | 0.0222 | -0.2025 | .84 | 1.0 |
| Total dwell time | 0.0139 | 0.0177 | 0.7865 | .44 | 1.0 |
| Group: Alzheimer's disease (n=19) | -4.9630 | 1.2990 | -3.8207 | <.001 | .003 |
| Interaction: Alzheimer's disease and total dwell time | 0.0083 | 0.0271 | 0.3074 | .76 | 1.0 |

The other HMMs provided comparable results.

*Table S7: Results from linear regression models predicting PANSS (sub)scores in participants with schizophrenia (n=18). PANSS_PT: Positive Scale, PANSS_NT: Negative Scale, PANSS_GT: General Psychopathology Scale, PANSS_CS: Composite Scale.*

| Response | Predictor | Coefficient | Standard error | *t* value | *P* value |
|---|---|---|---|---|---|
| PANSS_PT | Total dwell time | -0.0227 | 0.0543 | -0.4184 | .68 |
| | Age | -0.0015 | 0.1484 | -0.0098 | .99 |
| PANSS_NT | Total dwell time | 0.0018 | 0.0999 | 0.0176 | .99 |
| | Age | -0.0876 | 0.2731 | -0.3208 | .75 |
| PANSS_GT | Total dwell time | 0.0688 | 0.0954 | 0.7205 | .48 |
| | Age | 0.1898 | 0.2610 | 0.7273 | .48 |
| PANSS_CS | Total dwell time | -0.0245 | 0.0812 | -0.3013 | .77 |
| | Age | 0.0861 | 0.2220 | 0.3880 | .70 |
| PANSS_TOTAL | Total dwell time | 0.0478 | 0.2196 | 0.2178 | .83 |
| | Age | 0.1008 | 0.6005 | 0.1678 | .87 |

The other HMMs provided comparable results.

### Figures

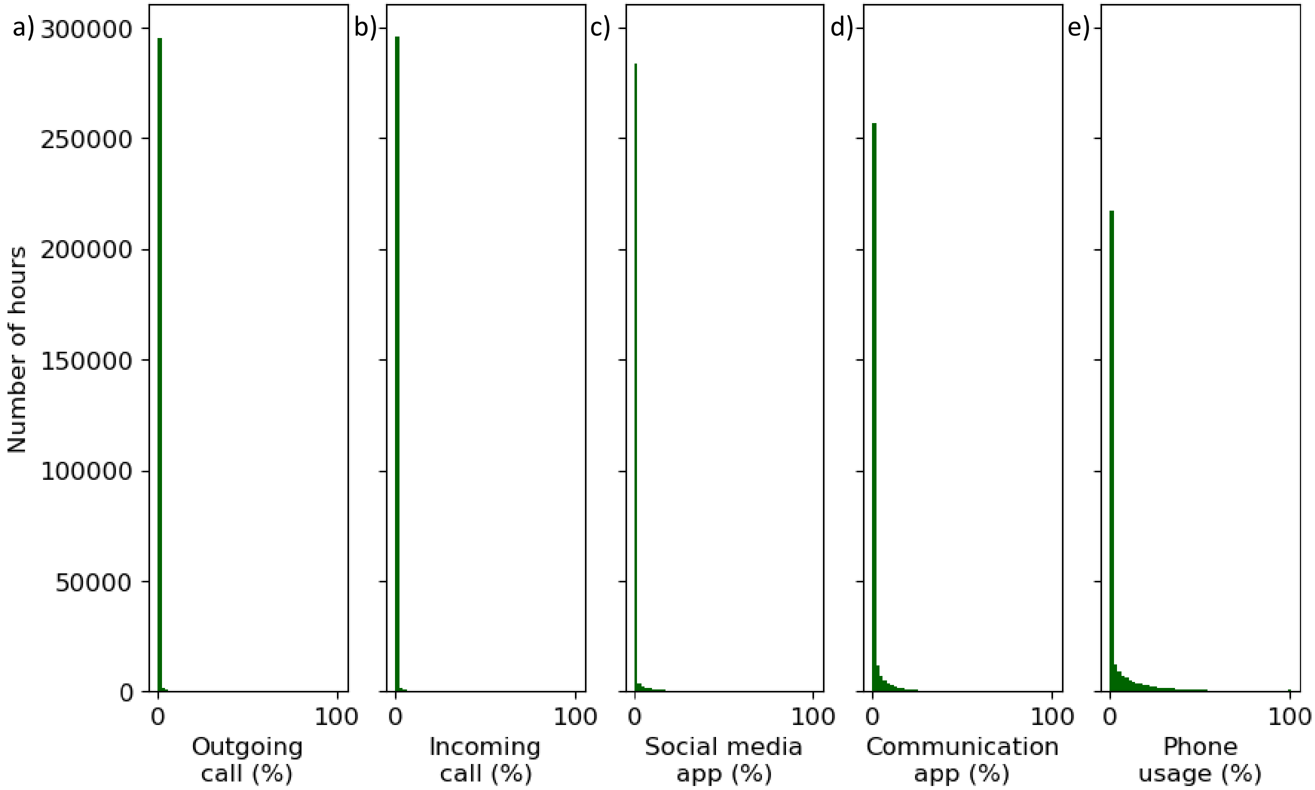


*Figure S1: Distributions of hourly percentage values per channel for all time series. It can be seen that these distributions are very zero-inflated. a) Outgoing call, b) incoming call, c) social media app, d) communication app, e) overall phone usage.*

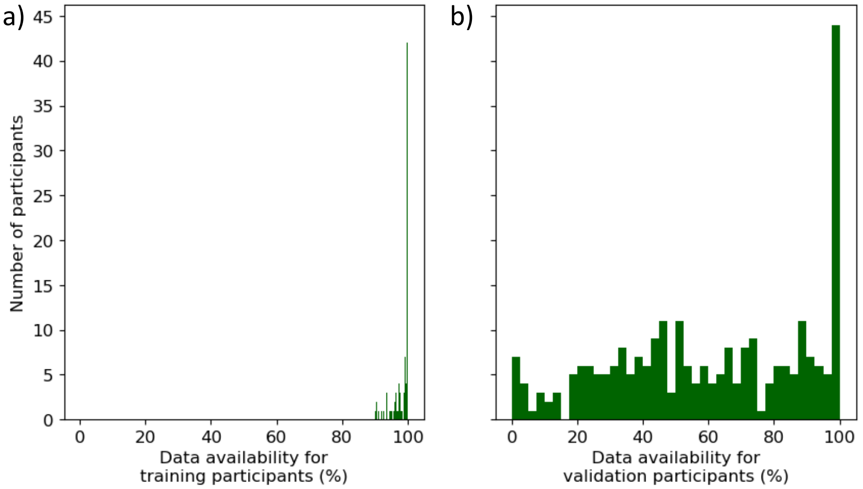


*Figure S2: Percentage of overall data availability for all participants, split by set. These values indicate missingness. a) Training participants, b) validation participants.*

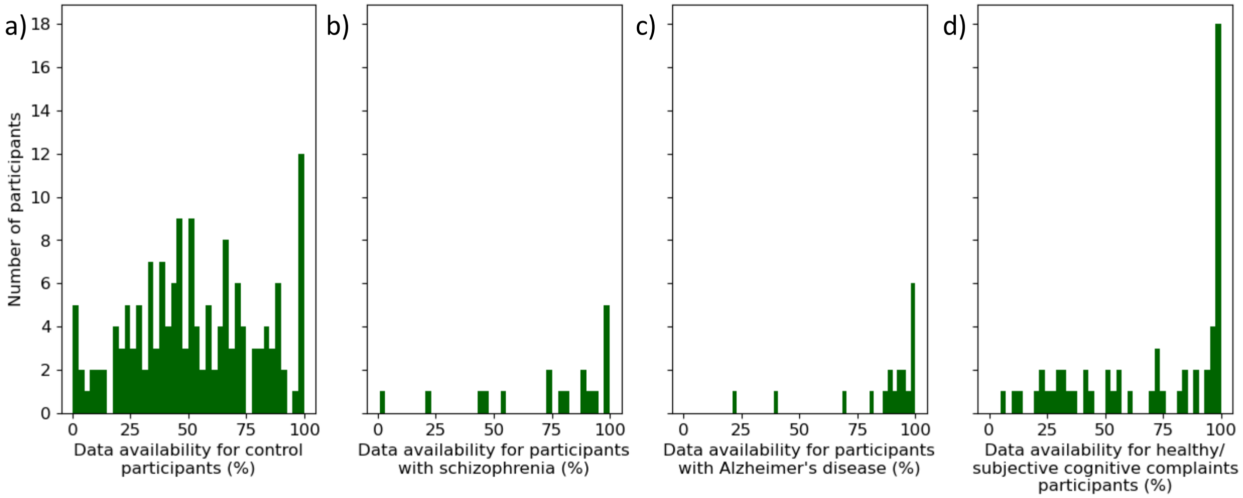


*Figure S3: Percentage of overall data availability for validation participants, split by diagnostic group. These values indicate missingness. a) Healthy controls, b) participants with schizophrenia, c) participants with Alzheimer's disease, d) subjective cognitive complaints participants.*

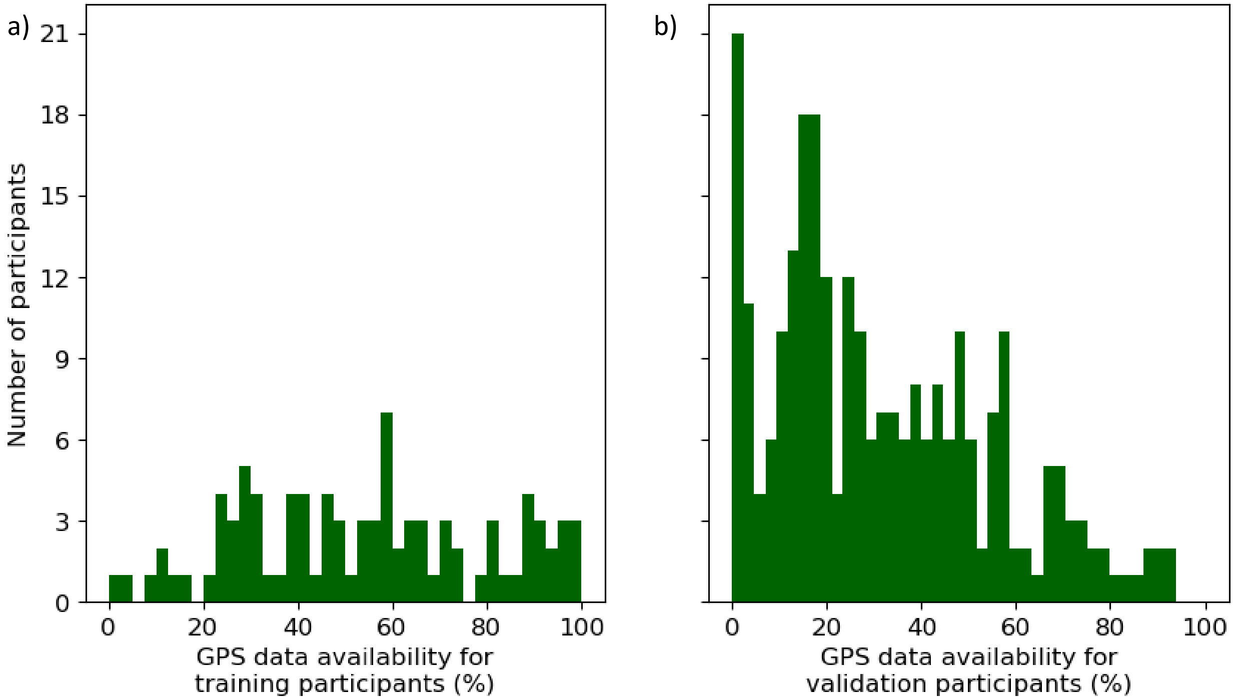


*Figure S4: Percentage of GPS data availability for all participants, split by set. These values indicate missing GPS data. a) Training participants, b) validation participants.*

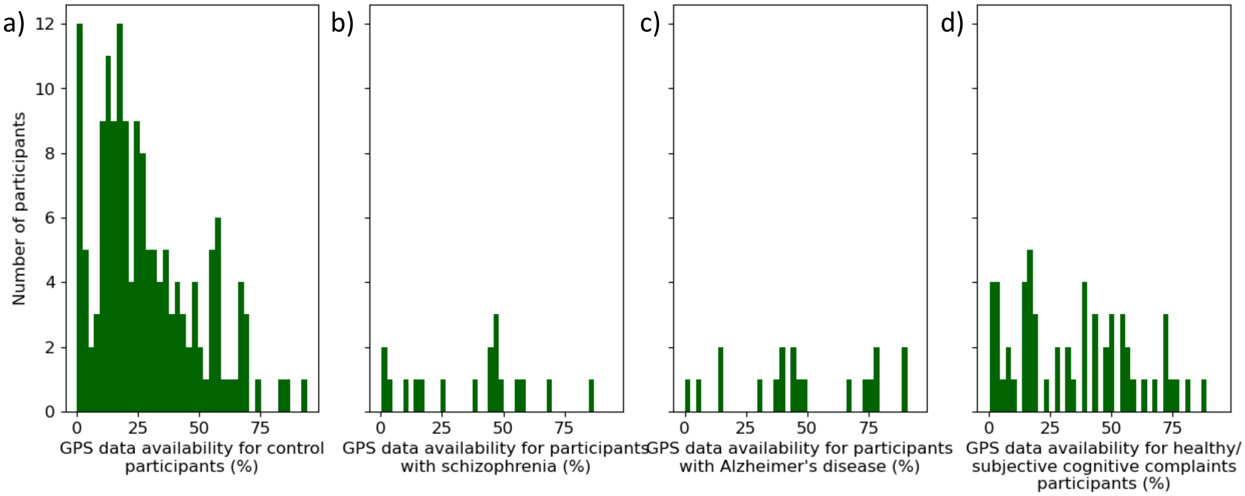


*Figure S5: Percentage of GPS data availability for validation participants, split by diagnostic group. These values indicate missing GPS data. a) Healthy controls, b) participants with schizophrenia, c) participants with Alzheimer's disease, d) subjective cognitive complaints participants.*

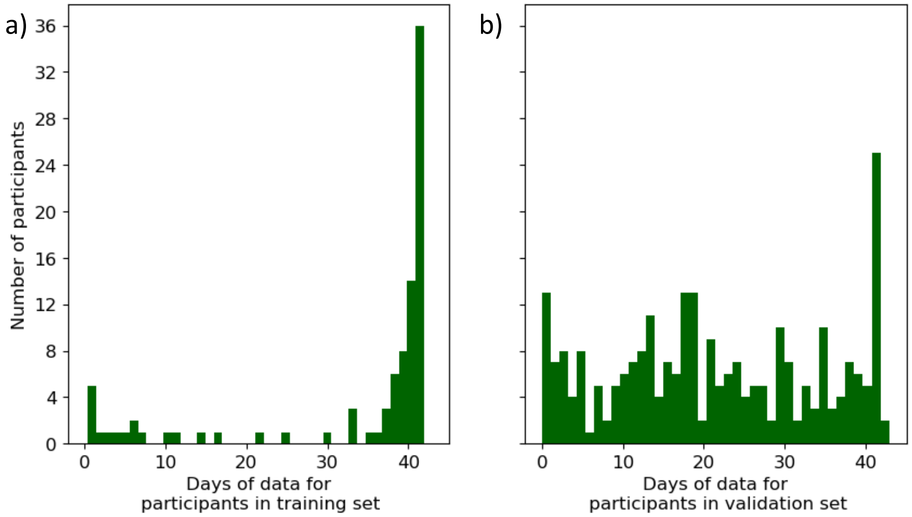


*Figure S6: Days of data for all participants, split by set. These values show the time series length. a) Training participants, b) validation participants.*

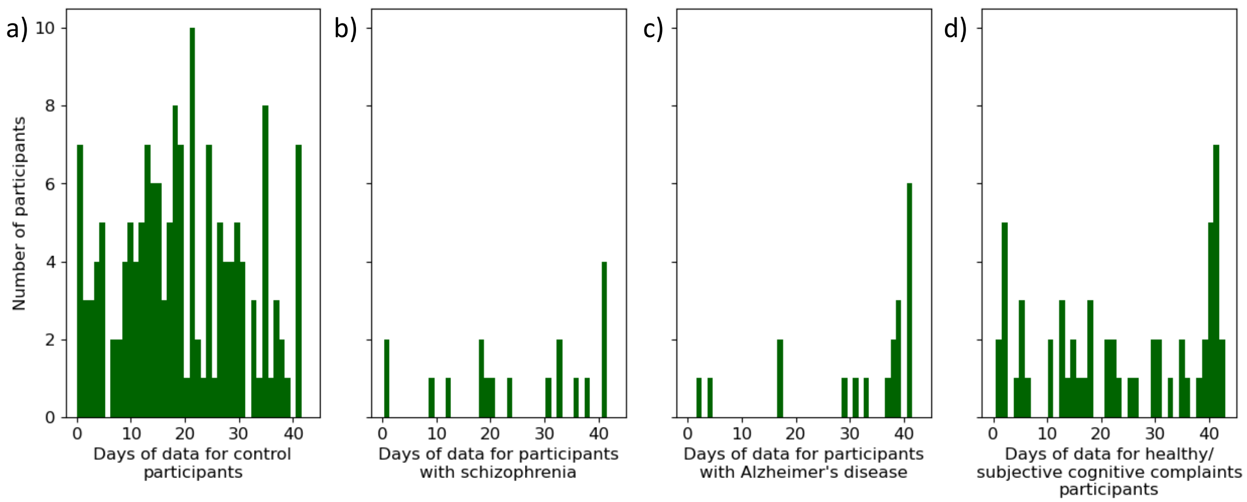


*Figure S7: Days of data for validation participants, split by diagnostic group. These values show the time series length. a) Healthy controls, b) participants with schizophrenia, c) participants with Alzheimer's disease, d) subjective cognitive complaints participants.*

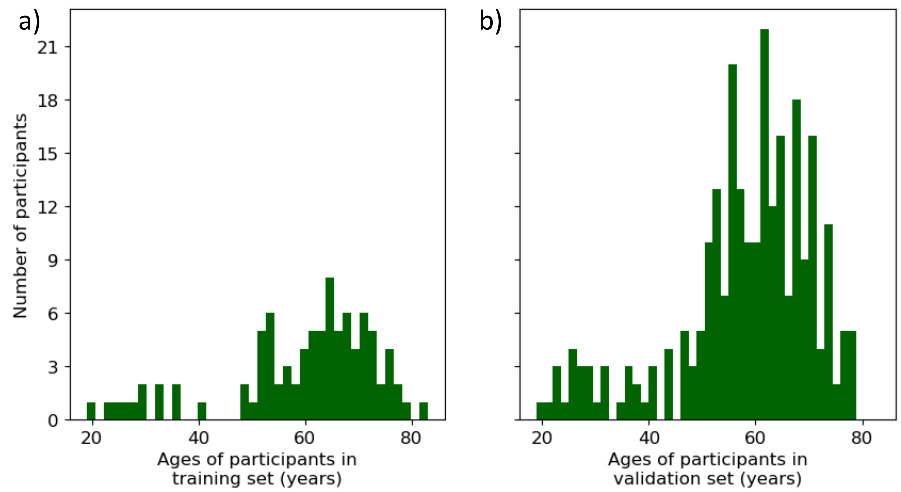


*Figure S8: Age distributions of all participants, split by set. a) Training participants, b) validation participants.*

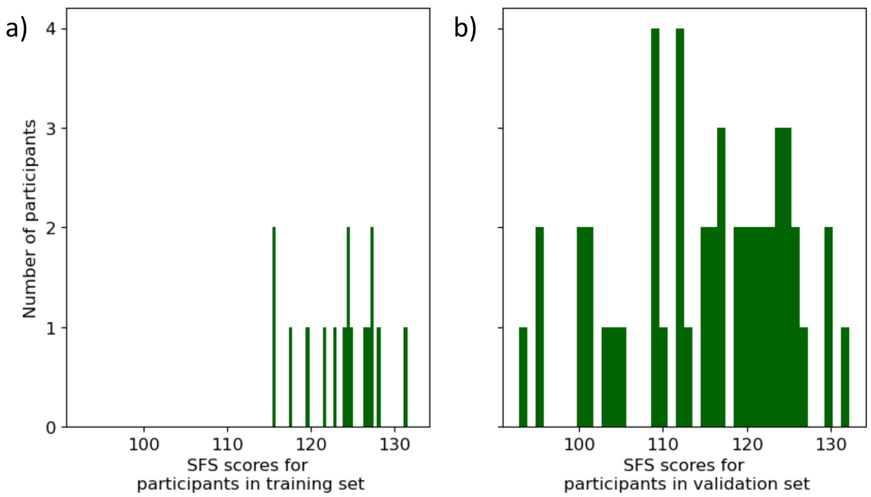


*Figure S9: Social Functioning Scale scores of all available participants, split by set. a) Training participants, b) validation participants. Possible range in scores: 55-135.*

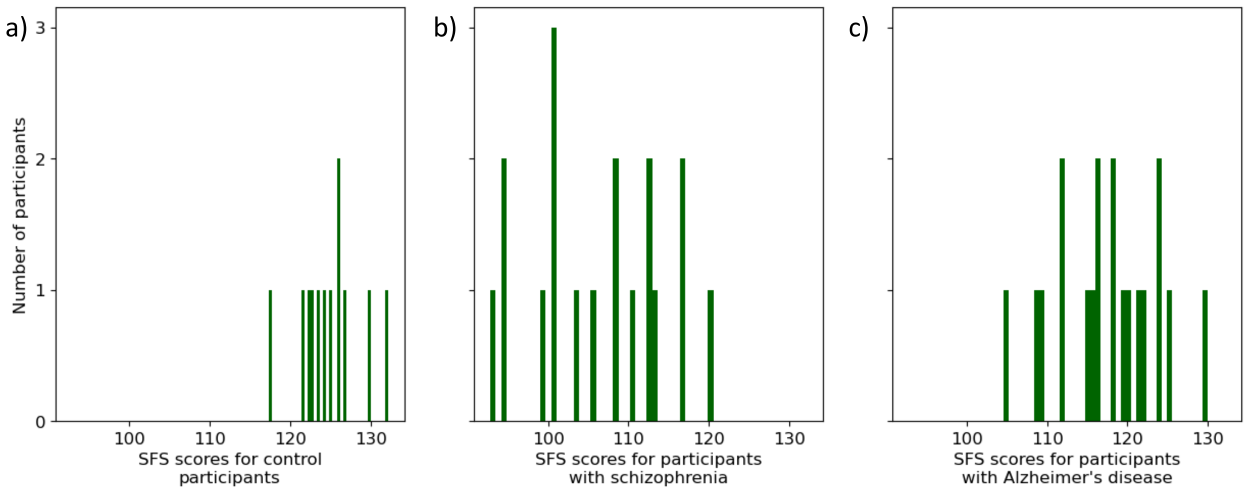


*Figure S10: Social Functioning Scale scores of available validation participants, split by diagnostic group. a) Healthy controls, b) participants with schizophrenia, c) participants with Alzheimer's disease.*

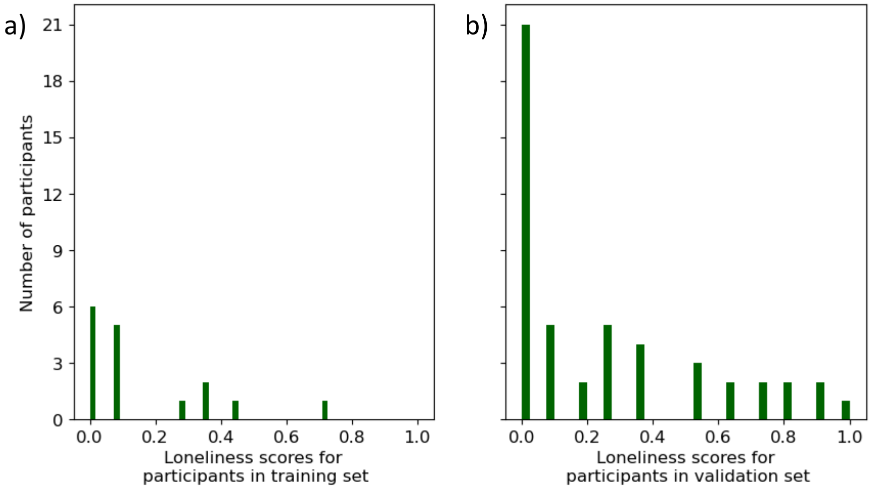


*Figure S11: Loneliness scores of all available participants, split by set. a) Training participants, b) validation participants. Possible range in scores: 0-11.*

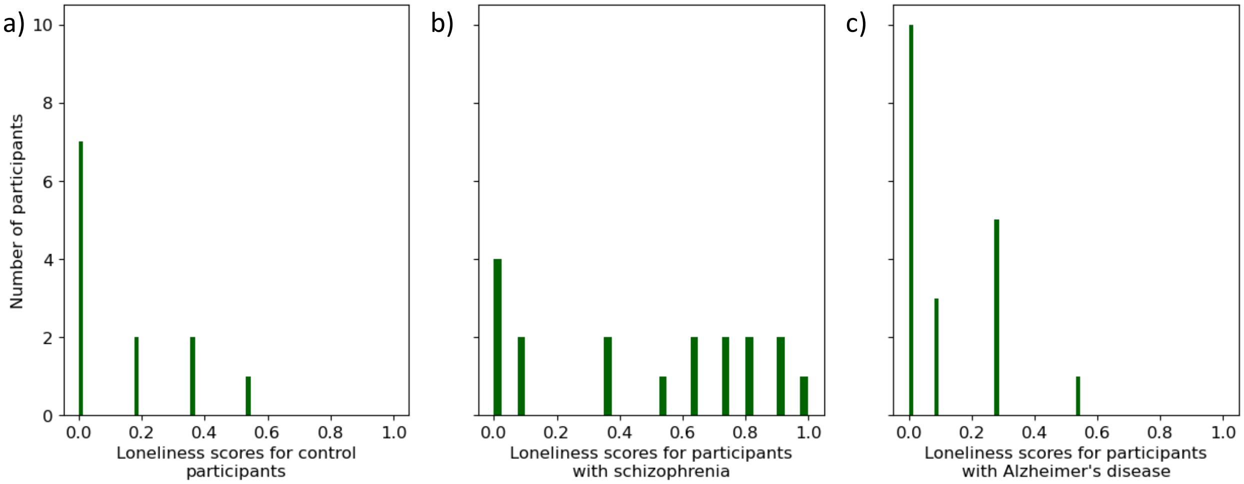


*Figure S12: Loneliness scores of available validation participants, split by diagnostic group. a) Healthy controls, b) participants with schizophrenia, c) participants with Alzheimer's disease.*

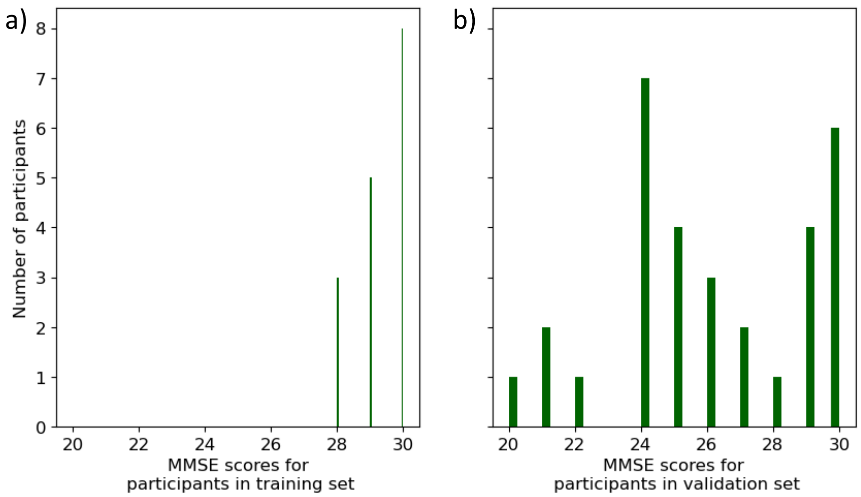


*Figure S13: Mini-Mental State Examination scores of all available participants, split by set. a) Training participants, b) validation participants. Maximum score = 30.*

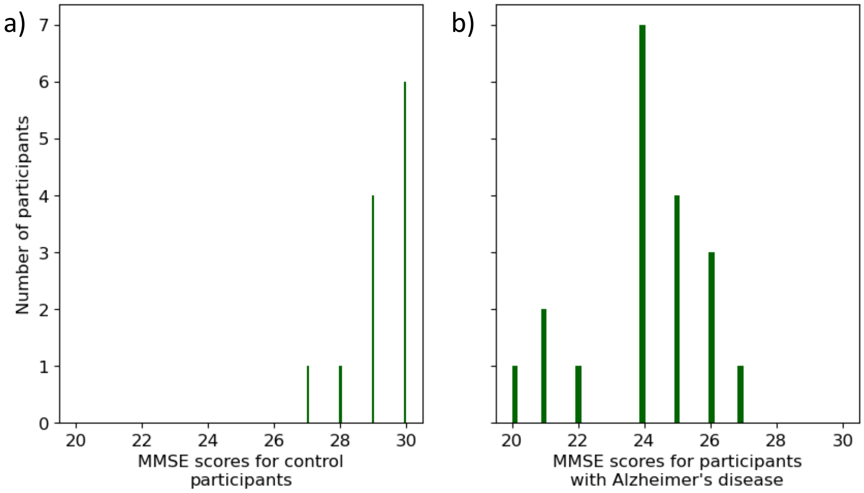


*Figure S14: Mini-Mental State Examination scores of available validation participants, split by diagnostic group. a) Healthy controls, b) participants with Alzheimer's disease.*

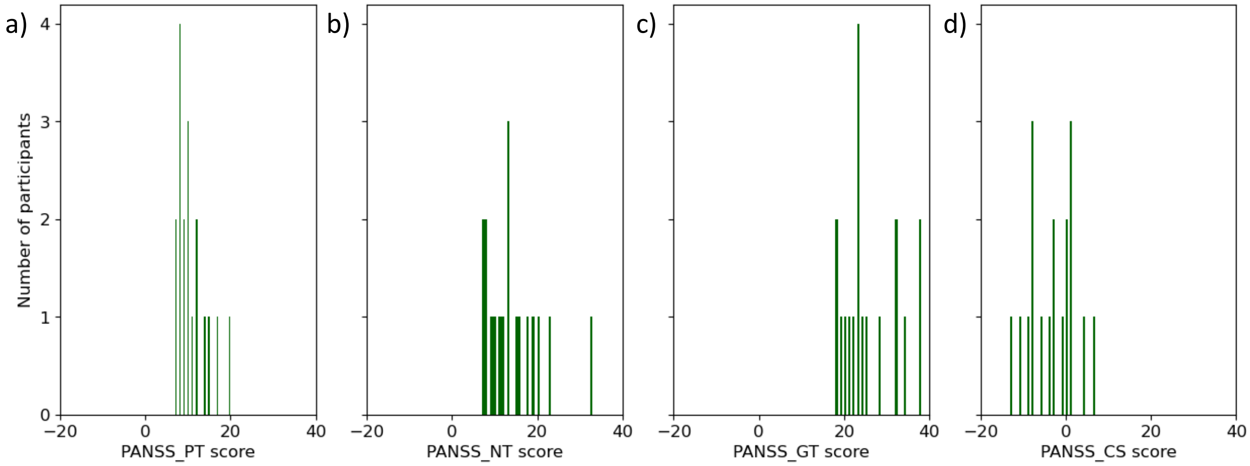


*Figure S15: Positive and Negative Syndrome Scale scores of participants with schizophrenia, split by (sub)scale. a) Positive symptom scores (PT), b) negative symptom scores (NT), c) general psychopathology scores (GT), d) composite scale scores (CS). Positive and Negative Scales range: 7-49; General Psychopathology Scale range: 16-112; Total PANSS score: sum of the sub scales; Composite Scale range: –42 to +42 (subtract negative score from positive score).*
