## Supplementary material for "Uncovering Social States in Healthy and Clinical Populations Using Digital Phenotyping and Hidden Markov Models: Observational Study": Multimedia Appendix 2. Supplementary Materials

### General information regarding additional HMMs

All of the relevant results for each HMM variation are reported one-by-one. For conciseness, we report the results from the state/s that were interpreted as socially active based on each HMM's emission probabilities, and where informative include a note of the results from statistical models from the other states.

#### Tables

*Table S1: In-sample Bayesian Information Criteria (BIC) for the different HMM variations investigated.*

| Data used for training | Number of states | Hour included as covariate? | In-sample BIC |
|---|---|---|---|
| Training set | 2 | No | 198580 |
| Training set | 3 | No | 195740 |
| Training set | 4 | No | 194315 |
| Training set | 2 | Yes | 188240 |
| Training set | 3 | Yes | 185295 |
| Training set | 4 | Yes | 185881 |
| All | 2 | No | 607748 |
| All | 3 | No | 594881 |
| All | 4 | No | 575001 |
| All | 2 | Yes | 586338 |
| All | 3 | Yes | 570134 |
| All | 4 | Yes | 548748 |

*SFS*

*Table S2: Three state HMM trained using training set, including hour as a covariate - results from a linear regression model predicting SFS score from total dwell time (state 1), age and group, where withheld healthy controls (n=12) were the reference group.*

| Predictor | Coefficient | Standard error | *t* value | *P* value | FDR corrected *P* value |
|---|---|---|---|---|---|
| Age | 0.0311 | 0.0809 | 0.3840 | .70 | 1.0 |
| Group: Schizophrenia (n=18) | -19.8194 | 6.2281 | -3.1823 | .003 | .02 |
| Group: Alzheimer's disease (n=19) | 3.9336 | 4.6803 | 0.8405 | .41 | 1.0 |
| Total dwell time | 0.1251 | 0.0692 | 1.8073 | .08 | .47 |
| Interaction: Schizophrenia and total dwell time | 0.0302 | 0.1057 | 0.2853 | .78 | 1.0 |
| Interaction: Alzheimer's disease and total dwell time | -0.3940 | 0.1248 | -3.1572 | .003 | .02 |

No significant interaction between Alzheimer’s disease group and total state 2 dwell time was identified. A significant interaction between Alzheimer’s disease group and total state 3 dwell time was identified.

*Table S3: Four state HMM trained using training set, including hour as a covariate - results from a linear regression model predicting SFS score from total dwell time (state 4), age and group, where withheld healthy controls (n=12) were the reference group.*

| Predictor | Coefficient | Standard error | *t* value | *P* value | FDR corrected *P* value |
|---|---|---|---|---|---|
| Age | 0.0335 | 0.0828 | 0.4049 | .69 | .83 |
| Group: Schizophrenia (n=18) | -18.7364 | 6.1746 | -3.0344 | .004 | .01 |
| Group: Alzheimer's disease (n=19) | 3.4459 | 4.6335 | 0.7437 | .46 | .69 |
| Total dwell time | 0.1210 | 0.0676 | 1.7892 | .08 | .16 |
| Interaction: Schizophrenia and total dwell time | 0.0099 | 0.1018 | 0.0972 | .92 | .92 |
| Interaction: Alzheimer's disease and total dwell time | -0.3781 | 0.1232 | -3.0681 | .004 | .01 |

*Table S4: Two state HMM trained using all data, including hour as a covariate - results from a linear regression model predicting SFS score from total dwell time (state 2), age and group, where all healthy controls (n=28) were the reference group.*

| Predictor | Coefficient | Standard error | *t* value | *P* value | FDR corrected *P* value |
|---|---|---|---|---|---|
| Age | 0.0662 | 0.0542 | 1.2219 | .23 | 1.0 |
| Group: Schizophrenia (n=18) | -21.2049 | 5.8024 | -3.6545 | <.001 | 0.003 |
| Group: Alzheimer's disease (n=19) | 2.4022 | 3.8025 | 0.6318 | .53 | 1.0 |
| Total dwell time | 0.0891 | 0.0530 | 1.6796 | .10 | 0.59 |
| Interaction: Schizophrenia and total dwell time | 0.0734 | 0.0947 | 0.7751 | .44 | 1.0 |
| Interaction: Alzheimer's disease and total dwell time | -0.2813 | 0.0893 | -3.1503 | .003 | 0.02 |

*Table S5: Three state HMM trained using all data, including hour as a covariate - results from a linear regression model predicting SFS score from total dwell time (state 1), age and group, where all healthy controls (n=28) were the reference group.*

| Predictor | Coefficient | Standard error | *t* value | *P* value | FDR corrected *P* value |
|---|---|---|---|---|---|
| Age | 0.0650 | 0.0552 | 1.1777 | .24 | 1.0 |
| Group: Schizophrenia (n=18) | -22.7680 | 5.4685 | -4.1635 | <.001 | <.001 |
| Group: Alzheimer's disease (n=19) | 1.9859 | 3.6657 | 0.5418 | .59 | 1.0 |
| Total dwell time | 0.0830 | 0.0534 | 1.5538 | .13 | 0.75 |
| Interaction: Schizophrenia and total dwell time | 0.1113 | 0.0951 | 1.1706 | .25 | 1.0 |
| Interaction: Alzheimer's disease and total dwell time | -0.3488 | 0.1109 | -3.1444 | .003 | 0.02 |

A significant interaction between Alzheimer’s disease and total state 2 dwell time was also identified. No significant interaction between Alzheimer’s disease and total state 3 dwell time was identified.

*Table S6: Four state HMM trained using all data, including hour as a covariate - results from a linear regression model predicting SFS score from total dwell time (state 2), age and group, where all healthy controls (n=28) were the reference group.*

| Predictor | Coefficient | Standard error | *t* value | *P* value | FDR corrected *P* value |
|---|---|---|---|---|---|
| Age | 0.0062 | 0.0481 | 0.1283 | .90 | .99 |
| Group: Schizophrenia (n=18) | -9.0947 | 2.6567 | -3.4234 | .001 | .003 |
| Group: Alzheimer's disease (n=19) | 0.0227 | 3.1312 | 0.0073 | .99 | .99 |
| Total dwell time | 0.0761 | 0.0449 | 1.6935 | .10 | .14 |
| Interaction: Schizophrenia and total dwell time | -0.3583 | 0.0850 | -4.2144 | <.001 | <.001 |
| Interaction: Alzheimer's disease and total dwell time | -0.2499 | 0.0838 | -2.9810 | .004 | .008 |

*Table S7: Four state HMM trained using all data, including hour as a covariate - results from a linear regression model predicting SFS score from total dwell time (state 4), age and group, where all healthy controls (n=28) were the reference group.*

| Predictor | Coefficient | Standard error | *t* value | *P* value | FDR corrected *P* value |
|---|---|---|---|---|---|
| Age | 0.0288 | 0.0612 | 0.4706 | .64 | .77 |
| Group: Schizophrenia (n=18) | -24.4169 | 3.0941 | -7.8914 | <.001 | <.001 |
| Group: Alzheimer's disease (n=19) | -6.8078 | 2.0879 | -3.2605 | .002 | .006 |
| Total dwell time | -0.0023 | 0.0468 | -0.0500 | .96 | .96 |
| Interaction: Schizophrenia and total dwell time | 0.2095 | 0.0697 | 3.0067 | .004 | .008 |
| Interaction: Alzheimer's disease and total dwell time | -0.2527 | 0.2141 | -1.1805 | .24 | .36 |

*Diagnostic group*

*Table S8: Three state HMM trained using training set, including hour as a covariate - results from a multinomial logistic regression model predicting diagnostic group (versus withheld healthy controls (n=156)) using total dwell time (state 1).*

| Group | Predictor | Coefficient | Standard error | Odds | *z* value | *P* value | FDR corrected *P* value |
|---|---|---|---|---|---|---|---|
| Schizophrenia (n=18) | Total dwell time | -0.0115 | 0.0171 | 0.9886 | -0.6711 | .50 | 1.0 |
| | Age | -0.1671 | 0.0306 | 0.8461 | -5.4614 | <.001 | <.001 |
| Alzheimer's disease (n=26) | Total dwell time | -0.0618 | 0.0148 | 0.9401 | -4.1698 | <.001 | <.001 |
| | Age | 0.0789 | 0.0295 | 1.0821 | 2.6716 | .008 | .02 |
| Subjective cognitive complaints (n=57) | Total dwell time | -0.0152 | 0.0084 | 0.9849 | -1.8116 | .07 | .21 |
| | Age | 0.0152 | 0.0165 | 1.0153 | 0.9192 | .36 | 1.0 |

The total state 2 dwell time was not a significant predictor of any of the diagnostic groups. Total state 3 dwell time was a significant predictor of Alzheimer's disease group and subjective cognitive complaints group.

*Table S9: Three state HMM trained using training set, including hour as a covariate - Binomial logistic regression age sensitivity analysis results (for state 1). A model was run per diagnostic group, where the reference group for each model was age-matched healthy controls. Age cut-offs for healthy controls (HC) matched to each diagnostic group (schizophrenia (SZ), Alzheimer's disease (AD), subjective cognitive complaints (SCC)): HC vs SZ, Age < 42 (HC n=12); HC vs AD, Age > 50 (HC n=130); HC vs SCC, Age > 43 (HC n=142).*

| Group | Predictor | Coefficient | Standard error | Odds | *z* value | *P* value | FDR corrected *P* value |
|---|---|---|---|---|---|---|---|
| Schizophrenia (n=18) | Total dwell time | -0.0060 | 0.0194 | 0.9940 | -0.3099 | .76 | 1.0 |
| | Age | 0.0605 | 0.0638 | 1.0624 | 0.9488 | .34 | 1.0 |
| Alzheimer's disease (n=26) | Total dwell time | -0.0641 | 0.0156 | 0.9380 | -4.1054 | <.001 | <.001 |
| | Age | 0.0518 | 0.0337 | 1.0531 | 1.5389 | .12 | .37 |
| Subjective cognitive | Total dwell time | -0.0148 | 0.0083 | 0.9853 | -1.7831 | .07 | .22 |

| | | | | | | | |
|---|---|---|---|---|---|---|---|
| complaints (n=57) | Age | -0.0133 | 0.0207 | 0.9868 | -0.6409 | .52 | 1.0 |

*Table S10: Four state HMM trained using training set, including hour as a covariate - results from a multinomial logistic regression model predicting diagnostic group (versus withheld healthy controls (n=156)) using total dwell time (state 4).*

| Group | Predictor | Coefficient | Standard error | Odds | *z* value | *P* value | FDR corrected *P* value |
|---|---|---|---|---|---|---|---|
| Schizophrenia (n=18) | Total dwell time | -0.0141 | 0.0166 | 0.9860 | -0.8461 | .40 | 1.0 |
| | Age | -0.1687 | 0.0307 | 0.8448 | -5.4909 | <.001 | <.001 |
| Alzheimer's disease (n=26) | Total dwell time | -0.0579 | 0.0142 | 0.9437 | -4.0775 | <.001 | <.001 |
| | Age | 0.0752 | 0.0297 | 1.0781 | 2.5295 | .01 | .03 |
| Subjective cognitive complaints (n=57) | Total dwell time | -0.0186 | 0.0083 | 0.9815 | -2.2462 | .02 | .07 |
| | Age | 0.0126 | 0.0166 | 1.0127 | 0.7617 | .45 | 1.0 |

*Table S11: Four state HMM trained using training set, including hour as a covariate - Binomial logistic regression age sensitivity analysis results (for state 4). A model was run per diagnostic group, where the reference group for each model was age-matched healthy controls. Age cut-offs for healthy controls (HC) matched to each diagnostic group (schizophrenia (SZ), Alzheimer's disease (AD), subjective cognitive complaints (SCC)): HC vs SZ, Age < 42 (HC n=12); HC vs AD, Age > 50 (HC n=130); HC vs SCC, Age > 43 (HC n=142).*

| Group | Predictor | Coefficient | Standard error | Odds | *z* value | *P* value | FDR corrected *P* value |
|---|---|---|---|---|---|---|---|
| Schizophrenia (n=18) | Total dwell time | -0.0054 | 0.0185 | 0.9946 | -0.2915 | .77 | 1.0 |
| | Age | 0.0602 | 0.0640 | 1.0621 | 0.9420 | .35 | 1.0 |
| Alzheimer's disease (n=26) | Total dwell time | -0.0587 | 0.0148 | 0.9430 | -3.9704 | <.001 | <.001 |
| | Age | 0.0503 | 0.0334 | 1.0516 | 1.5077 | .13 | .39 |
| Subjective cognitive complaints (n=57) | Total dwell time | -0.0192 | 0.0084 | 0.9810 | -2.2887 | .02 | .07 |
| | Age | -0.0176 | 0.0211 | 0.9826 | -0.8359 | .40 | 1.0 |

*Table S12: Two state HMM trained using all data, including hour as a covariate - results from a multinomial logistic regression model predicting diagnostic group (versus withheld healthy controls (n=247)) using total dwell time (state 2).*

| Group | Predictor | Coefficient | Standard error | Odds | *z* value | *P* value | FDR corrected *P* value |
|---|---|---|---|---|---|---|---|
| Schizophrenia (n=18) | Total dwell time | -0.0182 | 0.0164 | 0.9820 | -1.1087 | .27 | .80 |
| | Age | -0.1490 | 0.0258 | 0.8615 | -5.7663 | <.001 | <.001 |
| Alzheimer's disease (n=26) | Total dwell time | -0.0553 | 0.0131 | 0.9462 | -4.2351 | <.001 | <.001 |
| | Age | 0.0578 | 0.0266 | 1.0595 | 2.1678 | .03 | .09 |
| Subjective cognitive complaints (n=57) | Total dwell time | -0.0260 | 0.0082 | 0.9744 | -3.1518 | .002 | .005 |
| | Age | 0.0030 | 0.0141 | 1.0030 | 0.2162 | .83 | 1.0 |

*Table S13: Two state HMM trained using all data, including hour as a covariate - Binomial logistic regression age sensitivity analysis results (for state 2). A model was run per diagnostic group, where the reference group for each model was age-matched healthy controls. Age cut-offs for healthy controls (HC) matched to each diagnostic group (schizophrenia (SZ), Alzheimer's disease (AD), subjective cognitive complaints (SCC)): HC vs SZ, Age < 42 (HC n=24); HC vs AD, Age > 50 (HC n=206); HC vs SCC, Age > 43 (HC n=221).*

| Group | Predictor | Coefficient | Standard error | Odds | *z* value | *P* value | FDR corrected *P* value |
|---|---|---|---|---|---|---|---|
| Schizophrenia (n=18) | Total dwell time | -0.0126 | 0.0175 | 0.9875 | -0.7160 | .47 | 1.0 |
| | Age | 0.0423 | 0.0526 | 1.0432 | 0.8036 | .42 | 1.0 |
| Alzheimer's disease (n=26) | Total dwell time | -0.0557 | 0.0134 | 0.9458 | -4.1505 | <.001 | <.001 |
| | Age | 0.0298 | 0.0303 | 1.0302 | 0.9810 | .33 | .98 |
| Subjective cognitive complaints (n=57) | Total dwell time | -0.0283 | 0.0086 | 0.9721 | -3.3007 | <.001 | .003 |
| | Age | -0.0380 | 0.0196 | 0.9627 | -1.9371 | .05 | .16 |

*Table S14: Three state HMM trained using all data, including hour as a covariate - results from a multinomial logistic regression model predicting diagnostic group (versus withheld healthy controls (n=247)) using total dwell time (state 1).*

| Group | Predictor | Coefficient | Standard error | Odds | *z* value | *P* value | FDR corrected *P* value |
|---|---|---|---|---|---|---|---|
| Schizophrenia (n=18) | Total dwell time | -0.0211 | 0.0167 | 0.9791 | -1.2642 | .21 | .62 |
| | Age | -0.1529 | 0.0269 | 0.8582 | -5.6748 | <.001 | <.001 |
| Alzheimer's disease (n=26) | Total dwell time | -0.0647 | 0.0142 | 0.9373 | -4.5734 | <.001 | <.001 |
| | Age | 0.0540 | 0.0269 | 1.0555 | 2.0117 | .04 | .13 |
| Subjective cognitive complaints (n=57) | Total dwell time | -0.0187 | 0.0082 | 0.9815 | -2.2625 | .02 | .07 |
| | Age | 0.0045 | 0.0144 | 1.0045 | 0.3098 | .76 | 1.0 |

Total state 2 dwell time was a significant predictor of Alzheimer's disease group. However, total state 3 dwell time was not a significant predictor of any of the diagnostic groups.

*Table S15: Three state HMM trained using all data, including hour as a covariate - Binomial logistic regression age sensitivity analysis results (for state 1). A model was run per diagnostic group, where the reference group for each model was age-matched healthy controls. Age cut-offs for healthy controls (HC) matched to each diagnostic group (schizophrenia (SZ), Alzheimer's disease (AD), subjective cognitive complaints (SCC)): HC vs SZ, Age < 42 (HC n=24); HC vs AD, Age > 50 (HC n=206); HC vs SCC, Age > 43 (HC n=221).*

| Group | Predictor | Coefficient | Standard error | Odds | *z* value | *P* value | FDR corrected *P* value |
|---|---|---|---|---|---|---|---|
| Schizophrenia (n=18) | Total dwell time | -0.0187 | 0.0180 | 0.9815 | -1.0357 | .30 | .90 |
| | Age | 0.0386 | 0.0533 | 1.0394 | 0.7242 | .47 | 1.0 |
| Alzheimer's disease (n=26) | Total dwell time | -0.0663 | 0.0147 | 0.9358 | -4.5234 | <.001 | <.001 |
| | Age | 0.0239 | 0.0312 | 1.0241 | 0.7644 | .44 | 1.0 |
| Subjective cognitive complaints (n=57) | Total dwell time | -0.0184 | 0.0082 | 0.9818 | -2.2538 | .02 | .07 |
| | Age | -0.0333 | 0.0195 | 0.9673 | -1.7051 | .09 | .26 |

*Table S16: Four state HMM trained using all data, including hour as a covariate - results from a multinomial logistic regression model predicting diagnostic group (versus withheld healthy controls (n=247)) using total dwell time (state 2).*

| Group | Predictor | Coefficient | Standard error | Odds | *z* value | *P* value | FDR corrected *P* value |
|---|---|---|---|---|---|---|---|
| Schizophrenia (n=18) | Total dwell time | 0.0058 | 0.0134 | 1.0058 | 0.4362 | .66 | 1.0 |
| | Age | -0.1455 | 0.0262 | 0.8646 | -5.5527 | <.001 | <.001 |
| Alzheimer's disease (n=26) | Total dwell time | -0.0171 | 0.0114 | 0.9830 | -1.5030 | .13 | .40 |
| | Age | 0.0897 | 0.0259 | 1.0939 | 3.4649 | <.001 | .002 |
| Subjective cognitive complaints (n=57) | Total dwell time | -0.0152 | 0.0075 | 0.9849 | -2.0275 | .04 | .13 |
| | Age | 0.0195 | 0.0135 | 1.0197 | 1.4428 | .15 | .45 |

*Table S17: Four state HMM trained using all data, including hour as a covariate - Binomial logistic regression age sensitivity analysis results (for state 2). A model was run per diagnostic group, where the reference group for each model was age-matched healthy controls. Age cut-offs for healthy controls (HC) matched to each diagnostic group (schizophrenia (SZ), Alzheimer's disease (AD), subjective cognitive complaints (SCC)): HC vs SZ, Age < 42 (HC n=24); HC vs AD, Age > 50 (HC n=206); HC vs SCC, Age > 43 (HC n=221).*

| Group | Predictor | Coefficient | Standard error | Odds | *z* value | *P* value | FDR corrected *P* value |
|---|---|---|---|---|---|---|---|
| Schizophrenia (n=18) | Total dwell time | -0.0008 | 0.0145 | 0.9992 | -0.0567 | .95 | 1.0 |
| | Age | 0.0481 | 0.0537 | 1.0493 | 0.8958 | .37 | 1.0 |
| Alzheimer's disease (n=26) | Total dwell time | -0.0178 | 0.0114 | 0.9824 | -1.5618 | .12 | .36 |
| | Age | 0.0650 | 0.0282 | 1.0671 | 2.3005 | .02 | .06 |
| Subjective cognitive complaints (n=57) | Total dwell time | -0.0169 | 0.0075 | 0.9833 | -2.2377 | .03 | .08 |
| | Age | -0.0190 | 0.0188 | 0.9812 | -1.0138 | .31 | .93 |

*Table S18: Four state HMM trained using all data, including hour as a covariate - results from a multinomial logistic regression model predicting diagnostic group (versus withheld healthy controls (n=247)) using total dwell time (state 4).*

| Group | Predictor | Coefficient | Standard error | Odds | *z* value | *P* value | FDR corrected *P* value |
|---|---|---|---|---|---|---|---|
| Schizophrenia (n=18) | Total dwell time | -0.0157 | 0.0129 | 0.9845 | -1.2144 | .22 | .67 |
| | Age | -0.1556 | 0.0282 | 0.8559 | -5.5164 | <.001 | <.001 |
| Alzheimer's disease (n=26) | Total dwell time | -0.0817 | 0.0350 | 0.9216 | -2.3359 | .02 | .06 |
| | Age | 0.0650 | 0.0277 | 1.0672 | 2.3454 | .02 | .06 |
| Subjective cognitive complaints (n=57) | Total dwell time | -0.0059 | 0.0076 | 0.9941 | -0.7717 | .44 | 1.0 |
| | Age | 0.0114 | 0.0145 | 1.0115 | 0.7868 | .43 | 1.0 |

*Table S19: Four state HMM trained using all data, including hour as a covariate - Binomial logistic regression age sensitivity analysis results (for state 4). A model was run per diagnostic group, where the reference group for each model was age-matched healthy controls. Age cut-offs for healthy controls (HC) matched to each diagnostic group (schizophrenia (SZ), Alzheimer's disease (AD), subjective cognitive complaints (SCC)): HC vs SZ, Age < 42 (HC n=24); HC vs AD, Age > 50 (HC n=206); HC vs SCC, Age > 43 (HC n=221).*

| Group | Predictor | Coefficient | Standard error | Odds | *z* value | *P* value | FDR corrected *P* value |
|---|---|---|---|---|---|---|---|
| Schizophrenia (n=18) | Total dwell time | -0.0062 | 0.0126 | 0.9939 | -0.4863 | .63 | 1.0 |
| | Age | 0.0385 | 0.0549 | 1.0393 | 0.7017 | .48 | 1.0 |
| Alzheimer's disease (n=26) | Total dwell time | -0.0853 | 0.0361 | 0.9182 | -2.3630 | .02 | .05 |
| | Age | 0.0429 | 0.0309 | 1.0439 | 1.3886 | .16 | .49 |
| Subjective cognitive complaints (n=57) | Total dwell time | -0.0051 | 0.0077 | 0.9949 | -0.6645 | .51 | 1.0 |
| | Age | -0.0252 | 0.0195 | 0.9751 | -1.2930 | .20 | .59 |

### Figures

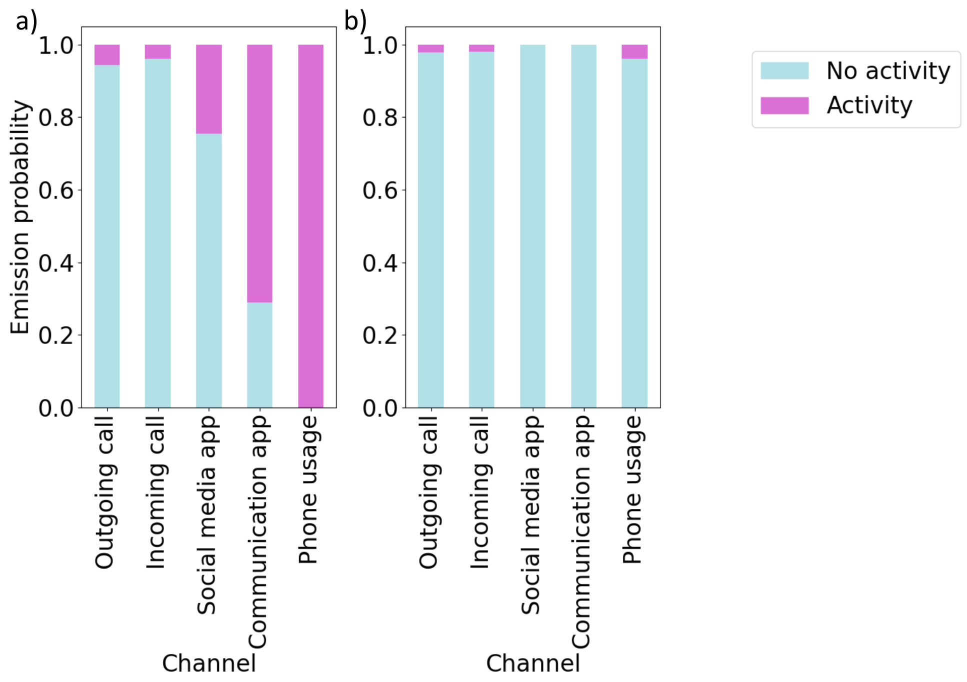


*Figure S1: Emission probabilities for two state HMM trained using all healthy controls, including hour as a covariate, for a) state 1 and b) state 2.*

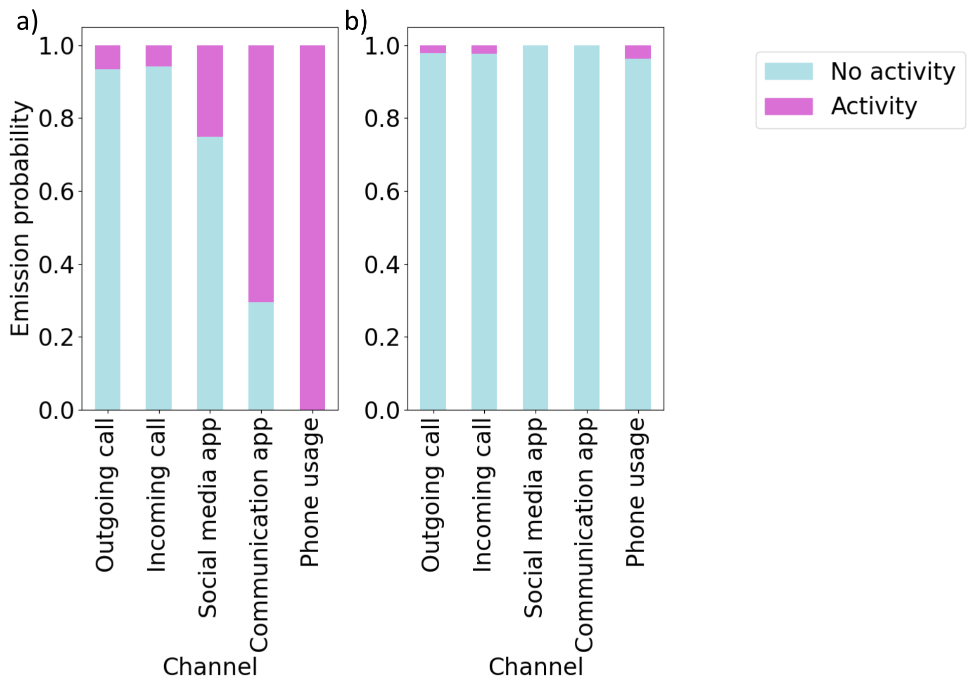


*Figure S2: Emission probabilities for two state HMM trained using all diagnostic groups, including hour as a covariate, for a) state 1 and b) state 2.*

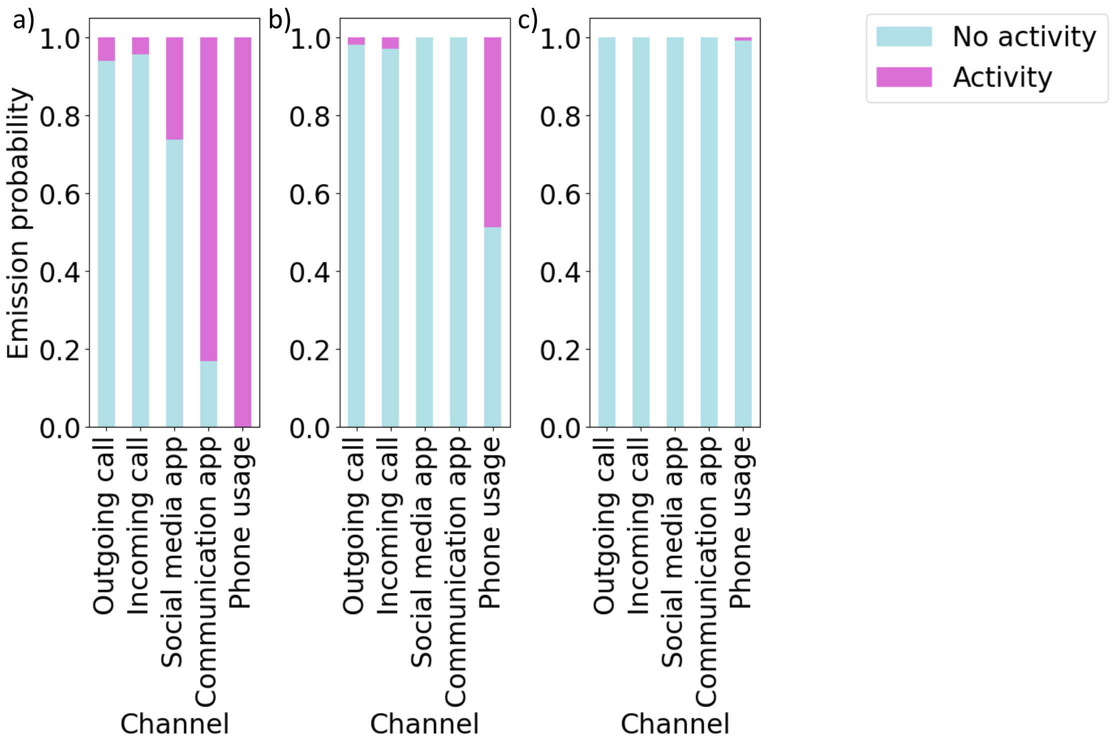


*Figure S3: Emission probabilities for three state HMM trained using training set, including hour as a covariate, for a) state 1, b) state 2 and c) state 3.*

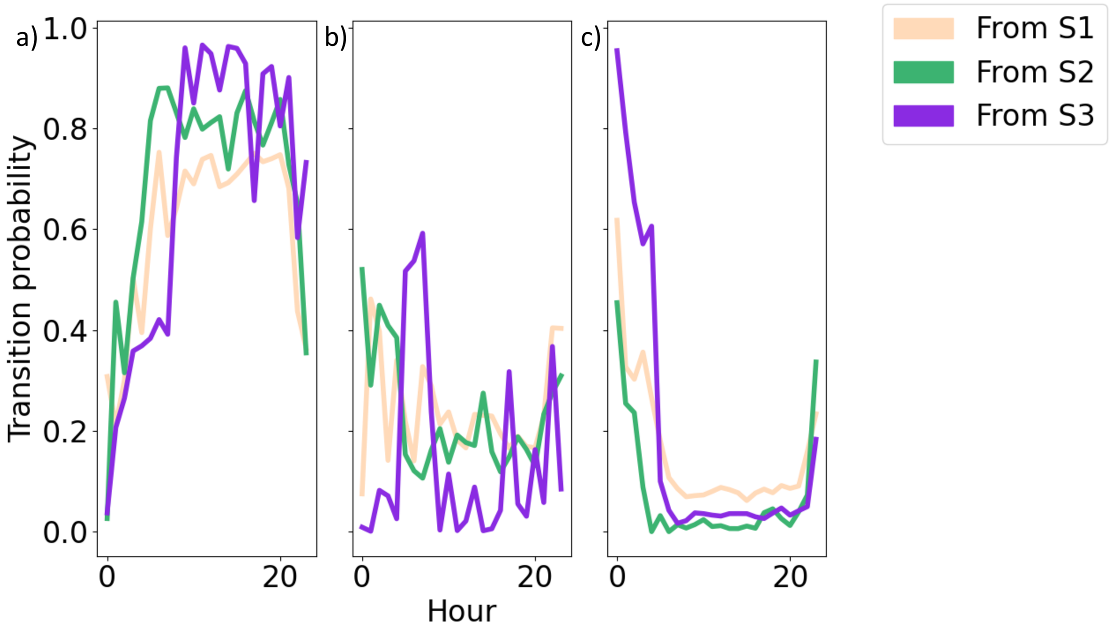


*Figure S4: Transition probabilities for three state HMM trained using training set, including hour as a covariate, reflecting transitions to a) state 1, b) state 2 and c) state 3.*

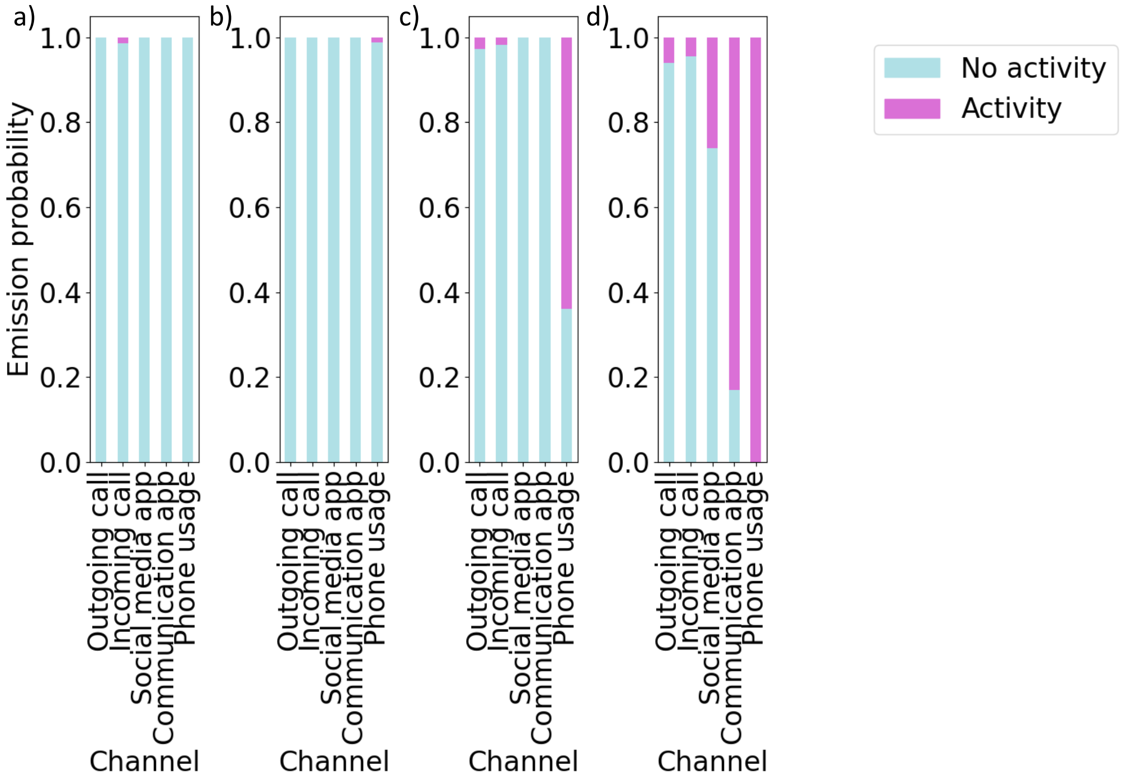


*Figure S5: Emission probabilities for four state HMM trained using training set, including hour as a covariate, for a) state 1, b) state 2, c) state 3 and d) state 4.*

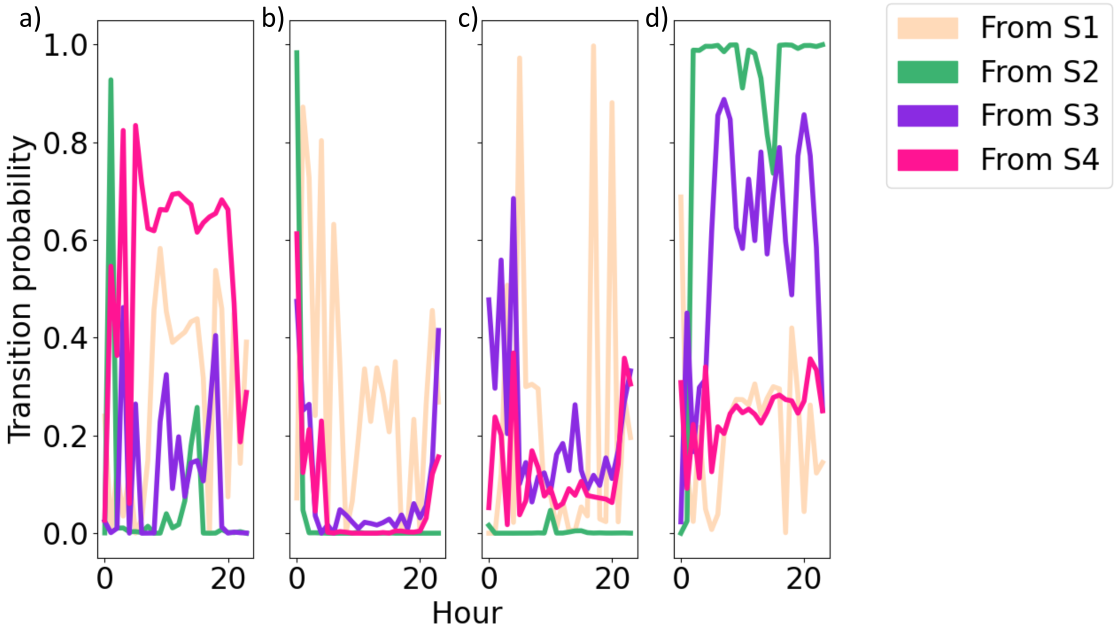


*Figure S6: Transition probabilities for four state HMM trained using training set, including hour as a covariate, reflecting transitions to a) state 1, b) state 2, c) state 3 and d) state 4.*

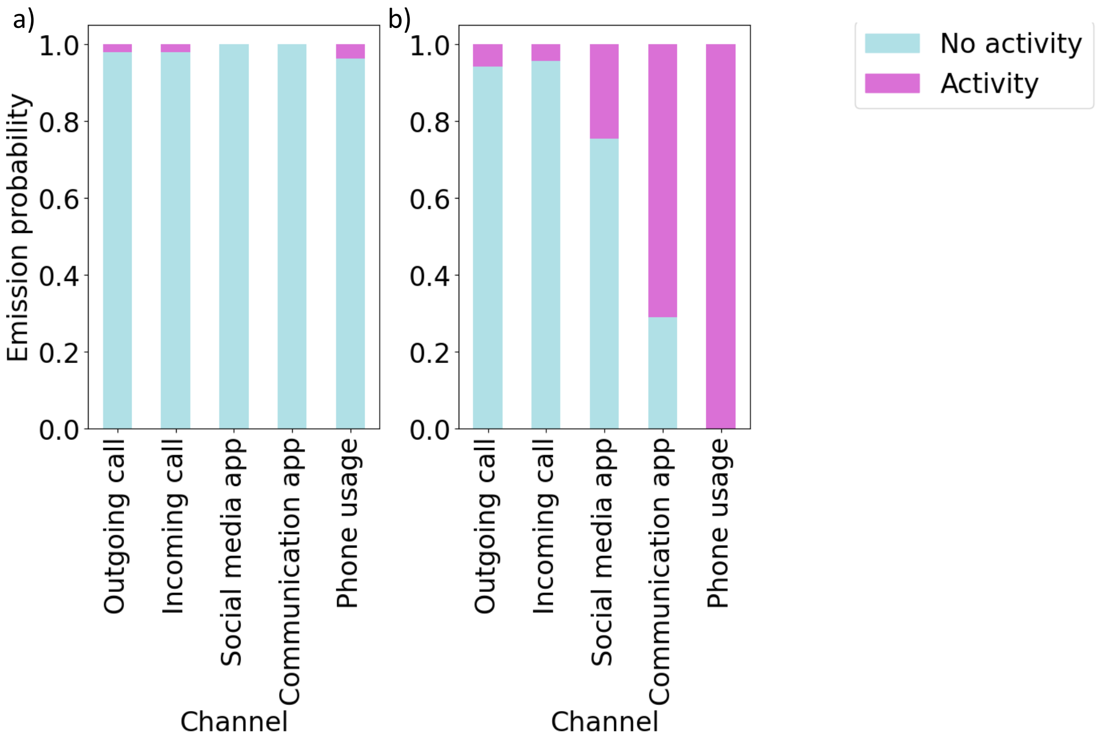


*Figure S7: Emission probabilities for two state HMM trained using all data, including hour as a covariate, for a) state 1 and b) state 2.*

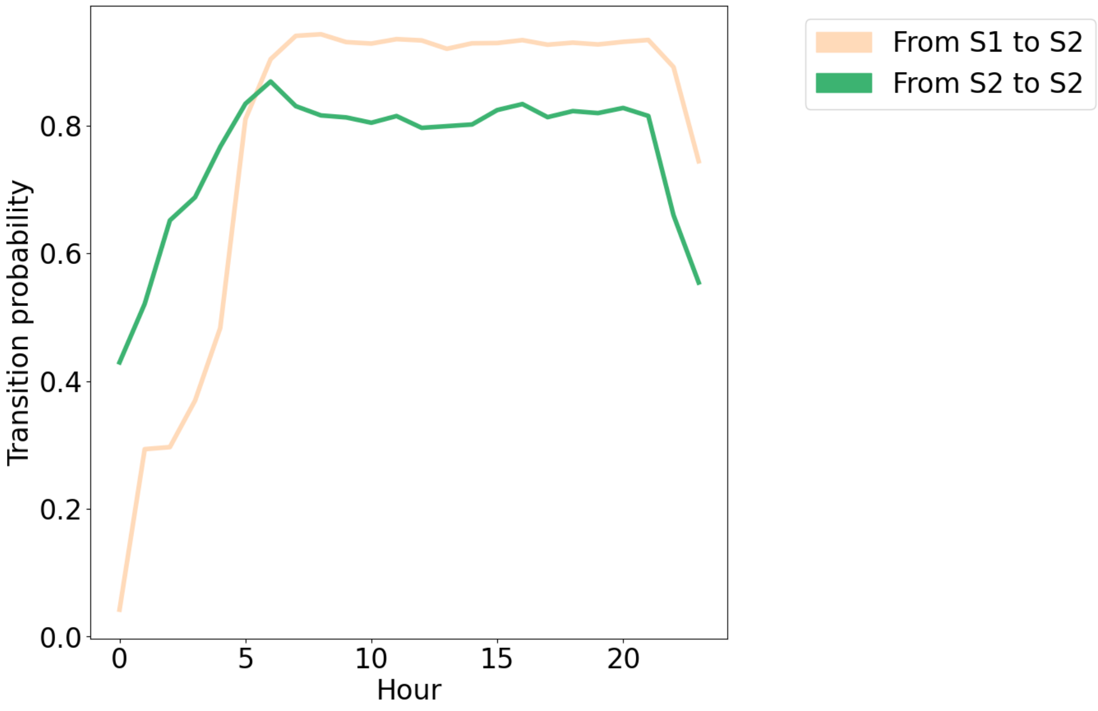


*Figure S8: Transition probabilities for two state HMM trained using all data, including hour as a covariate.*

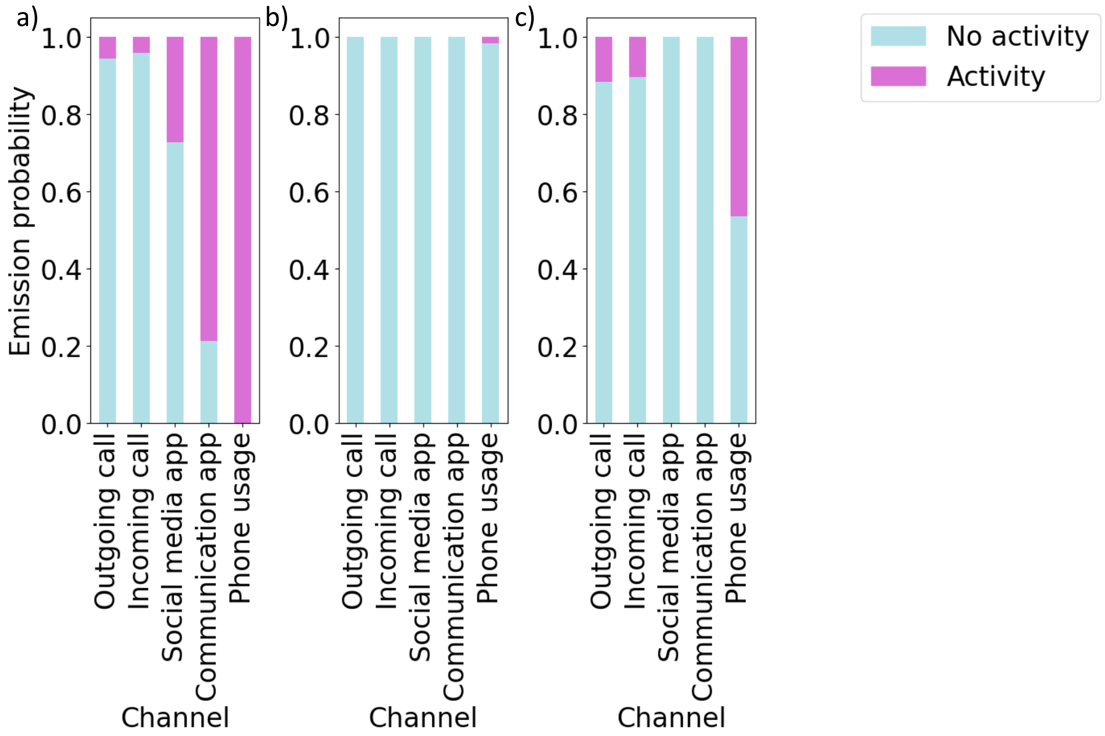


*Figure S9: Emission probabilities for three state HMM trained using all data, including hour as a covariate, for a) state 1, b) state 2 and c) state 3.*

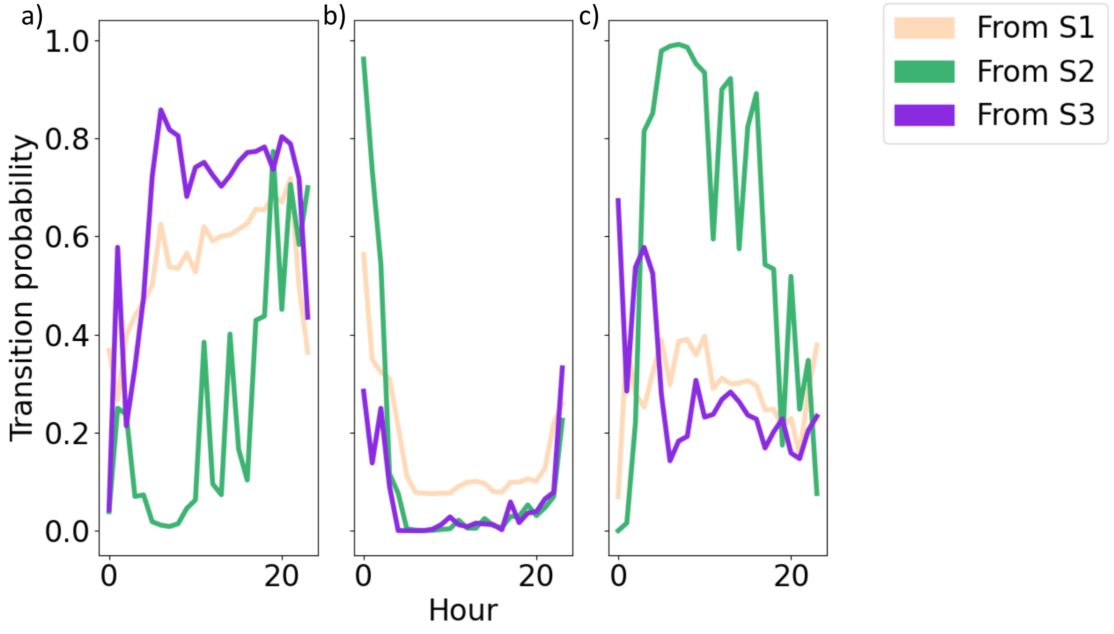


*Figure S10: Transition probabilities for three state HMM trained using all data, including hour as a covariate, reflecting transitions to a) state 1, b) state 2 and c) state 3.*

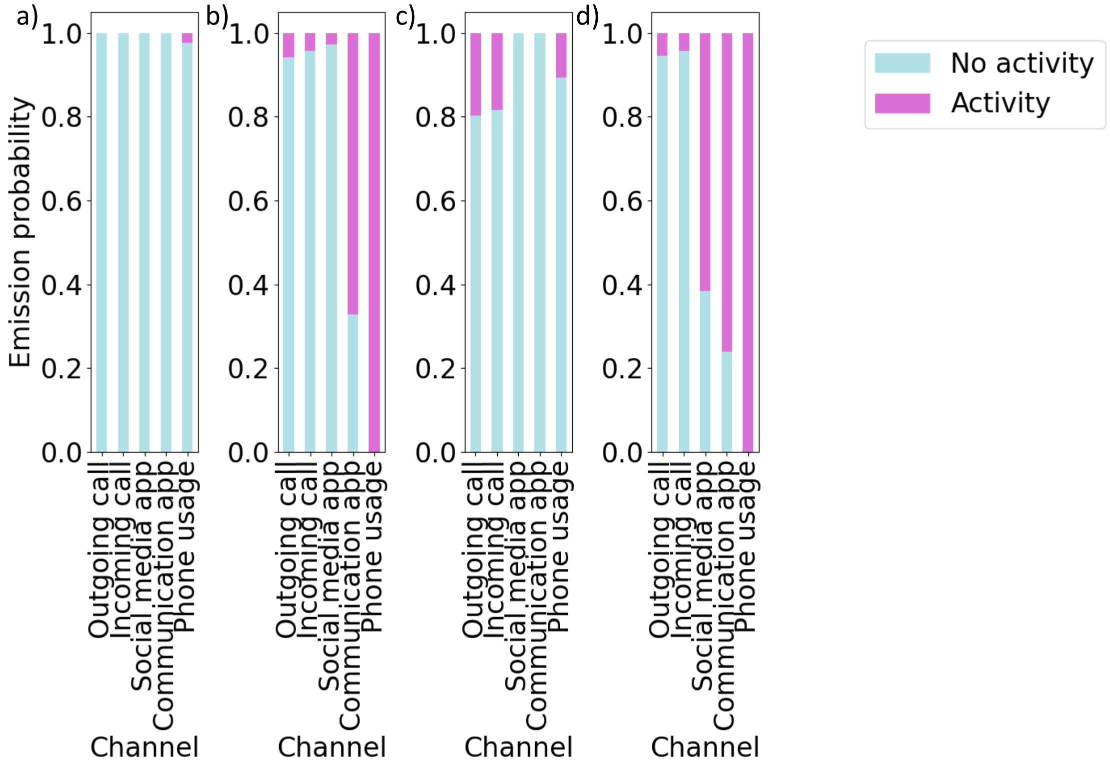


*Figure S11: Emission probabilities for four state HMM trained using all data, including hour as a covariate, for a) state 1, b) state 2, c) state 3 and d) state 4.*

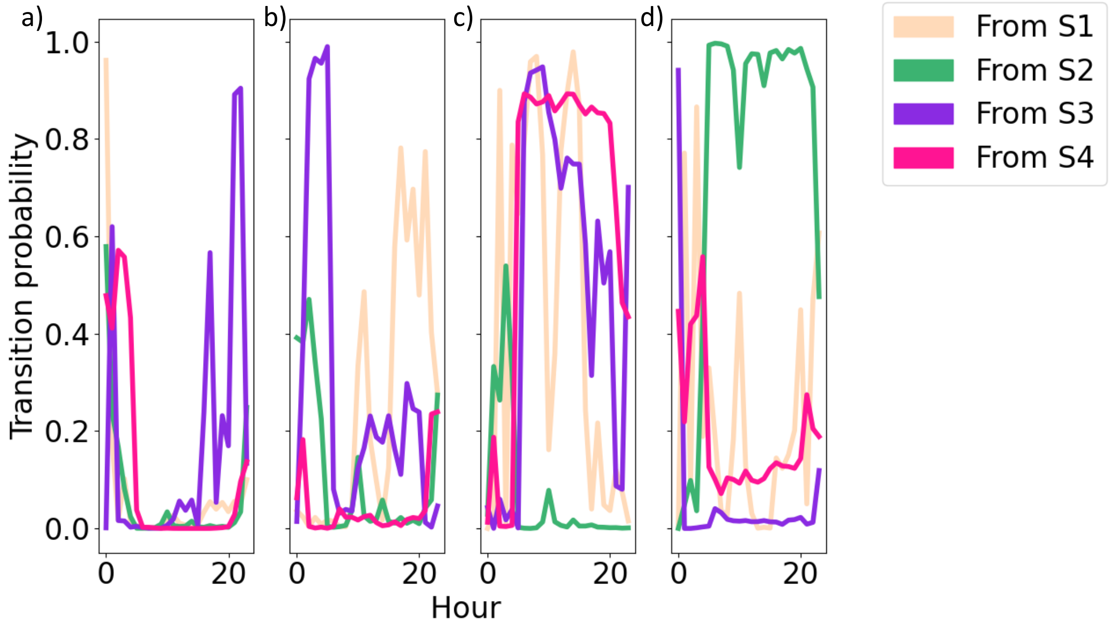


*Figure S12: Transition probabilities for four state HMM trained using all data, including hour as a covariate, reflecting transitions to a) state 1, b) state 2, c) state 3 and d) state 4.*
